# Caspases in COVID-19 Disease and Sequela and the Therapeutic Potential of Caspase Inhibitors

**DOI:** 10.1101/2020.11.02.20223636

**Authors:** Matthew Plassmeyer, Oral Alpan, Michael J. Corley, Kimberleigh Lillard, Paige Coatney, Tina Vaziri, Suzan Michalsky, Thomas A. Premeaux, Alina P.S. Pang, Zaheer Bukhari, Stephen T. Yueng, Teresa H Evering, Gail Naughton, Martin Latterich, Philip Mudd, Alfred Spada, Nicole Rindone, Denise Loizou, Lishomwa C. Ndhlovu, Raavi Gupta

**Author notes:** Corresponding Author: Lishomwa C. Ndhlovu MD, PhD, Professor-Elect, Department of Medicine, Division of Infectious Diseases, Weill Cornell Medicine, 413E 69^th^ St, New York, NY. Co-Senior Authors (Lishomwa C. Ndhlovu, Raavi Gupta).

## Abstract

Currently, there is no effective vaccine and only one FDA approved early-stage therapy against SARS-CoV-2 infection as an indication to prevent disease progression. Cellular caspases play a role in the pathophysiology of a number of disorders that the co-morbid conditions seen in severe COVID-19 disease. In this study, we assessed transcriptional states of caspases in blood cells from COVID-19 patients. Gene expression levels of select caspases were increased in *in vitro* SARS-CoV-2 infection models and single cell RNA-Seq data of blood from COVID-19 patients showed a distinct caspase expression in T cells, neutrophils, and dendritic cells. Flow cytometric evaluation of CD4 T cells showed up-regulation of caspase-1 in hospitalized COVID-19 patients compared to unexposed controls. Convalescent COVID-19 patients with lingering symptoms (“long haulers”) showed persistent up-regulation of caspase-1 in CD4 T cells that was attenuated *ex vivo* following co-culture with a select pan-caspase inhibitor. Further, we observed elevated caspase-3 levels in red blood cells from COVID-19 patients compared to controls that were responsive to caspase inhibition. Our results expose an exuberant caspase response in COVID-19 that may facilitate immune-related pathological processes leading to severe outcomes. Pan-caspase inhibition could emerge as a therapeutic strategy to ameliorate or prevent severe COVID-19 outcomes.

## Introduction

Coronavirus Disease 2019 (COVID-19) is the latest global health threat and, as in two preceding instances of the emergence of coronavirus respiratory disease, Severe Acute Respiratory Syndrome and Middle East Respiratory Syndrome, poses critical challenges for the public health, research, and medical communities (1, 2). While a robust research effort is currently under way to develop a vaccine against SARS-CoV-2, the causal agent of COVID-19, a variety of investigational therapeutic approaches are also being explored (3). Although the pathology of COVID-19 is now well described, the mechanisms of disease progression is still not clear. In clinical trials dexamethasone reduced severe outcomes in critically ill patients, suggesting of an inflammatory mechanism. However, while the use of specific anti-IL-6 monoclonal antibody therapies attenuated the cytokine storm, and eculizumab reduced soluble inflammatory markers, including c-reactive protein, in moderate-severe COVID-19, the clinical benefit has been marginal (4, 5). These findings suggest a lack of understanding of the effector molecules responsible for disease progression or that an intervention earlier in the course of the disease is needed, in order to help design effective therapies to ameliorate disease manifestations and its complications (6-8).

The scope and severity of COVID-19 varies among those infected, with some patients presenting with no or minor flu-like symptoms and quick recovery, some have sustained fever and have persistent fatigue with a post-viral syndrome, while others experience serious lung involvement that requires hospitalization and may lead to death (9). Although the respiratory and the gastrointestinal system are initial targets for SARS-CoV-2, there clearly is a systemic nature to this disease in some individuals that may be driven by micro-emboli and inflammatory processes (10, 11). Follow up in natural history studies will likely uncover additional post-infection sequelae. Furthermore, the notable impairment in type-I interferon responses and rapid lymphopenia clearly plays a role in disease severity (12-14). The scope and severity of COVID-19 is extraordinary, ranging from patients who present with no or minor flu-like symptoms and recover quickly to those who experience sustained fever and have persistent fatigue with a post-viral syndrome, to people who have serious lung involvement that either results in hospitalization or creates intubation needs and intensive care to people who die (14). This highlights the need for novel therapeutics that take into consideration the mechanism(s) of infection, viral replication, and effector pathways that lead to COVID-19 associated pathologies.

We recently reported that caspase-1 expression in lymphocytes and serum IL-18 levels are increased in liver transplant patients acutely ill with SARS-CoV-2 infection, along with non-liver transplant controls, suggesting pyroptosis mechanisms may play role in severe COVID-19 (15). A recent study showed that SARS-CoV-2 infection of rhesus macaques led to an upregulation of caspase-1 molecular signature in peripheral blood cells as early as day 2 post-inoculation (16). Pyroptosis, also known as caspase-1-dependent cell death, is inherently inflammatory, triggered by various pathological stimuli (i.e. stroke, heart attack, cancer), crucial for controlling microbial infections (17-19), and characterized by rapid plasma-membrane rupture and the release of pro-inflammatory intracellular contents (20, 21), a marked contrast to the regulated death process of apoptosis (22). Insight into the complex activation and regulation of the inflammasome complex and the way in which COVID-19 intersects with this pathway is an area of significant investigation (23). Thus, strategies targeting the inflammasome/pyroptosis pathway upstream of the production of the effector cytokines may be a novel approach to reverse COVID-19 induced immune perturbations (24). Building on our previous findings, we sought to expand our analysis to investigate the expression of not only the inflammatory caspases but also initiator and executionary caspases across the spectrum of COVID-19 disease in multiple immune cell types. The finding of increases in caspase molecules beyond caspase-1, such as caspase-3 in red blood cells (RBCs), led us to further define the full caspase expression profile of immune system cells. The impact of unique caspase expression profiles in given cells may impact specific outcomes such as RBCs involvement in coagulopathies in COVID-19 disease and determine the relationship with parameters of disease progression (25, 26).

## Materials and Methods

### Patient population

COVID-19 patient blood samples used for immunophenotyping were obtained during patients’ visit or hospitalizations at SUNY Downstate Medical Center in New York from May through to July 2020. Patients were defined as 1) non-hospitalized, with and without presentation of COVID-19 symptoms and 2) hospitalized with presentation of COVID-19 symptoms. Peripheral blood from venipuncture was drawn into EDTA and Heparin coated vacutainer tubes for immunophenotyping and processed within 48h of blood draw. Control blood samples from healthy volunteers without SARS CoV-2 infection or co-morbid conditions were collected after obtaining written informed consent.

### Flow Cytometry

Whole blood was stained per the clinical standard immunophenotyping protocols (Amerimmune LLC, Fairfax, VA). The samples were stained with the multiple antibody combinations for 30 minutes at 4°C. RBCs were lysed using BD FACS lysis solution (BD Bioscience, San Jose, CA) as per manufacture directions. In brief, freshly obtain peripheral blood mononuclear cells (PBMC) were separated from 2 mL of whole blood within 24h of collection and diluted 1:1 with phosphate buffered saline pH 7.2 (PBS) (Thermo Fisher Scientific, Carlsbad, CA) using Lymphoprep (Stem cell Technologies, Cambridge, MA) and Accuspin tubes (Sigma-Aldrich, St. Louis, MO) as per manufactures directions. PBMCs’ were washed in PBS and resuspended in 0.5 mL PBS. 100 μL of the PBMCs were immunostained with a mixture of antibodies at 4°C for 1 hour. Cells were washed and resuspended in PBS prior to acquisition. Antibodies used for the immune phenotyping of patient samples are detailed in the Supplementary information section of the manuscript (Supplemental Figure 1).

**Figure 1.**
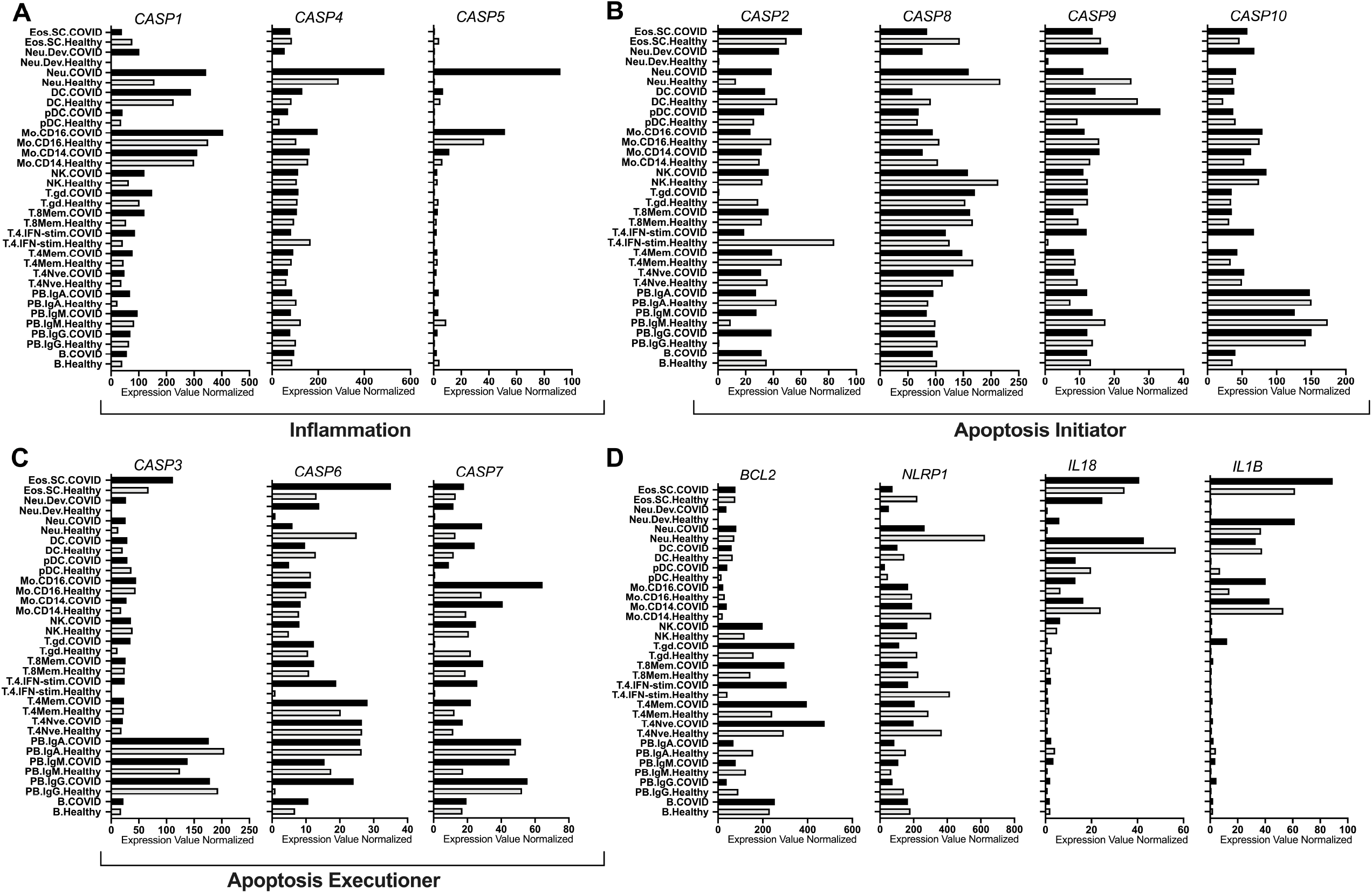
Human caspase single cell gene expression level. Data was compiled from three COVID-19 participants and from 6 healthy controls. Expression values for the caspase genes were normalized by DESeq2.

Apoptosis and pyroptosis were measured by flow cytometry using fluorescent-labeled inhibitors of caspase probe assay (FLICA; Immunochemistry Technologies, Minneapolis, MN). As a control, PBMCs were stimulated with nigericin for 2 hours. FAM-FLICA probes specific for caspase-1 were added to 50 μl PBMC and incubated for 1h at 37°C. Cells were subsequently washed and stained for CD45 PE-CY7 [HI30], CD3 AF700 [UCHT1], CD4 PE [RPA-T4], CD45RO PerCP-EF710 [UCHL1] and Viability Dye 780 (Thermo Fisher Scientific, Carlsbad, CA) Samples were acquired on a 3 laser BD FACS Canto 10. CS&T beads (BD Bioscience, San Jose, CA) were acquired daily to ensure consistent performance of the BD FACS Canto 10 Canto10. The BD FACS Canto 10 utilized for this study has been validated for T, B, NK and Dendritic cell immunophenotyping clinical diagnostic testing. Denovo FCS Express v6 clinical edition (De Novo Software, Pasadena, CA) was used for flow cytometric analyses.

### Plasma experiments

Plasma was separated from whole blood following centrifugation at 960 RCF. Cells (RBC and WBC) were either incubated at 37 °C alone or in the presence of trypsin for 1 hour then washed with 10 packed cell volumes of RPMI 1640 incomplete medium. Plasma was either held at room temperature (18 - 25°C) or heat inactivated at 56°C for 1 hr. Plasma was added back to the RBC/WBC in a 1:1 ratio and incubated overnight, rocking at room temperature.

### Public SARS-CoV-2 and COVID-19 Transcriptome Analyses

Single cell RNA-Seq data from three COVID-19 participants that were ventilated and diagnosed with acute respiratory distress syndrome at 2-16 days after symptom onset and from 6 healthy controls was accessed from GEO (27). RNA-Seq data from cell lines infected *in vitro* with SARS-CoV-2 was accessed from GEO: GSE147507 (28). Expression values for Caspase genes were normalized by DESeq2.

### Ex vivo stimulation studies

Active caspase-1 in COVID-19+ patient samples. Whole blood from a COVID-19 positive patient was either (A) untreated or (B) treated with the pan-caspase inhibitor emricasan (Sigma Aldrich, MO, SML2227-5MG) or selective caspase-1 inhibitor VX765 overnight at 37 degrees in a water bath. Subsequently, PBMCs were purified (Accuspin System – Histopaque 1077; Sigma Aldrich, MO, A6929) and incubated with nigericin (Immunochemistry Technologies, MN) for 2h. A Fam-FLICA probe specific for active caspase-1 was added to 50 μl PBMC, incubated for 1h at 37°C. PMBCs were washed with cell wash buffer (Immunochemistry Technologies, MN) and stained with CD45 PE-CY7 [HI30], CD3 AF700 [UCHT1], CD4 PE [RPA-T4], CD45RO PerCP-EF710 [UCHL1] and Viability Dye 780 (Thermo Fisher Scientific, Carlsbad, CA). Lymphocytes were identified using a standard gating schematic which incorporated gating of lymphocytes on an FSC/SSC plot and singlets on a FSC-A/FSC-H plot. Lymphocytes were further identified as CD45+ on a CD45/SSC plot and subsequent CD3-, CD3+ and CD3+CD4+ cells were identified on a CD45+ CD3/CD4 plot. Shown are the active caspase-1 MFI and % positive for the three cellular populations indicated.

### Statistical analysis

Demographic and HIV-related characteristics were described using the median, first quartile (Q1), and third quartile (Q3) for continuous variables and frequency for categorical variables. Differences among continuous variables were evaluated by either the Mann-Whitney test or Krustal-Wallis test with Dunn’s multiple comparisons. Relationships among parameters were examined by Pearson correlation for continuous variables. All statistical tests were performed with GraphPad Prism version 8.0 (Graphpad Software Inc., CA, USA). Statistical significance is indicated as *p<0.05, **p<0.01, ***p<0.001, ****p<0.0001. P-values ≤0.100, but not significant, are noted as statistical trends.

### Human studies Approval

All clinical investigation were conducted according to Declaration of Helsinki principles. All human studies were approved by institutional review boards (IRB 269846-10 and 1285028 protocols from State University of New York Downstate Medical Center and Amerimmune respectively). Written informed consent was received from participants prior to inclusion in the study.

## Results

### Transcriptional profile of multiple caspases in immune cells during COVID-19 disease

To follow up on our findings of increase caspase-1 expression in T cells of patients with COVID-19, we assayed for multiple caspases in different immune cell types from blood samples of patients with moderate-severe COVID-19. We examined caspase gene expression levels in public transcriptome profiling datasets of single cell RNA-Seq of immune cells from individuals with COVID-19 (Figure 1) and in *in vitro* SARS-CoV-2 infected cell (Supplemental Figures 1-3) and found transcriptional support of our previous findings showing upregulation of caspase-1 in CD4 T cells. We also found evidence of altered transcriptome levels of caspase genes in natural killer (NK cells), and neutrophils. Interestingly, plasmacytoid dendritic cells were the only immune cell type that showed up-regulation of caspase-9. Neutrophils showed a unique profile with upregulated caspase-5 and 7, an inflammatory and a pro-apoptotic caspase respectively. IFN-stimulated CD4 T cells show significant upregulation of caspase-7 and -9. We found that RNA-Seq data from several cell lines infected *in vitro* with SARS-CoV-2 showed an increase in caspase 1 and 4 in select cell lines relative to other viruses, suggesting multiple cellular death mechanisms that may potentially play a role in addition to the caspase-1 pathway.

### Changes in intracellular active Caspase 1 levels in immune cells in hospitalized patients with COVID-19 disease

We designed a laboratory developed test (LDT) to stain intracellular active caspase-1 in CD4 T cells and analytically validated it in a CLIA certified and CAP accredited flow cytometry laboratory. We analyzed intracellular levels of active caspase-1 in T cells and other immune cell types in COVID-19 participants (non-ICU and ICU) and healthy individuals for comparison (Figure 2A-D; demographics of participants are detailed in Supplementary Table 1). Frequency of caspase-1+ CD4 T cells were significantly elevated at baseline in hospitalized (both ICU and non-ICU) COVID-19 patients compared to healthy participants with and without nigericin stimulation (all p-value<0.0001).

**Figure 2.**
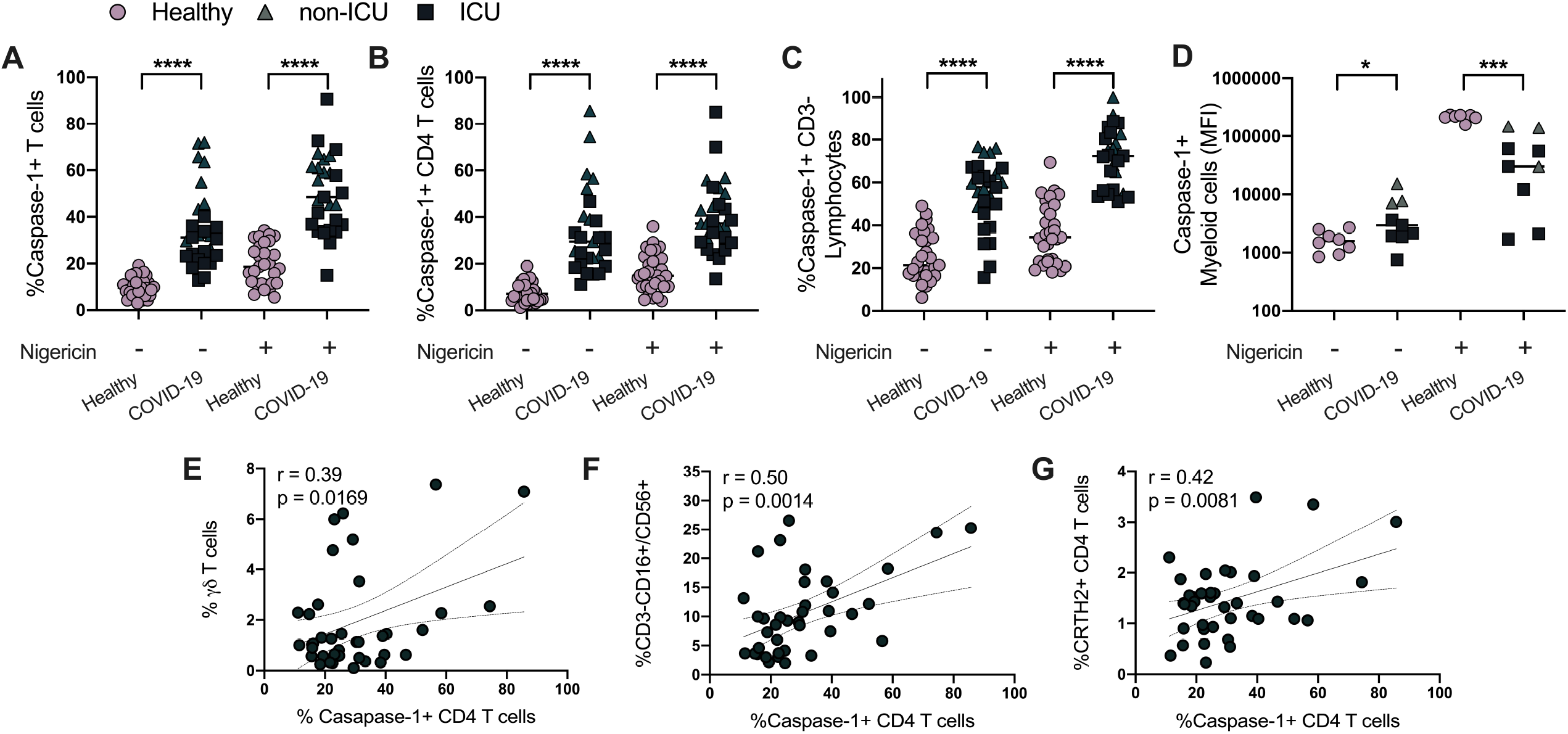
Caspase-1 expression in immune cells. Caspase-1 expression is shown for (**A-C)** Total (CD3+) T-cells, CD4 T-cells, non-CD3+ T cell lymphocytes, and myeloid cells. **(D-F)** Caspase 1 expression in CD4 T cells is correlated with *γ*δ T cells, NK cells (CD3-CD56/16+) and CRTH2+ CD4 T-cells.

Although we did not see increase in IL-18 and IL-1β in the RNA analysis, serum levels of IL-18 were increased in moderate-severe COVID-19 individuals and showed a positive correlation with T-helper cell caspase-1 expression (data not shown). Caspase-1 expression is predominantly in the CD45RO memory population and showed a weak but statistically significant correlation with older age, a finding that might potentially explain advanced age as one of the biggest risk factor for poor outcomes in COVID-19 (Supplemental Table 2). Furthermore, CD4 T cell caspase-1 levels in patients with COVID-19 correlated with CRTH2+ T-cells, *γ*/δ T-cells, CD3-CD16+/CD56+ lymphocytes, and plasmacytoid dendritic cells (Figure 1 E-G and Supplemental Table 2). Such correlations point out to the complex cellular interactions involved in COVID-19.

### Active Caspase-1 levels in CD4 T cells in non-COVID-19 patients

To determine if elevated caspase-1 levels in CD4 T cells is unique to COVID-19 patients, we assessed these levels in non-COVID-19 patients presenting to an Allergy/Immunology Clinic (Supplemental Table 3) and assays were performed as a part of patient care during their routine immunological work-up. Data from 102 adult and pediatric subjects are shown in Figure 2 for patients who presented with chronic sinusitis, moderate-severe asthma, chronic idiopathic urticaria and immune deficiencies. Normal ranges for the assay are shown in gray shaded areas (Figure 3). Only patients with asthma showed a baseline elevation of caspase-1 in CD4 T cells (Figure 3).

**Figure 3.**
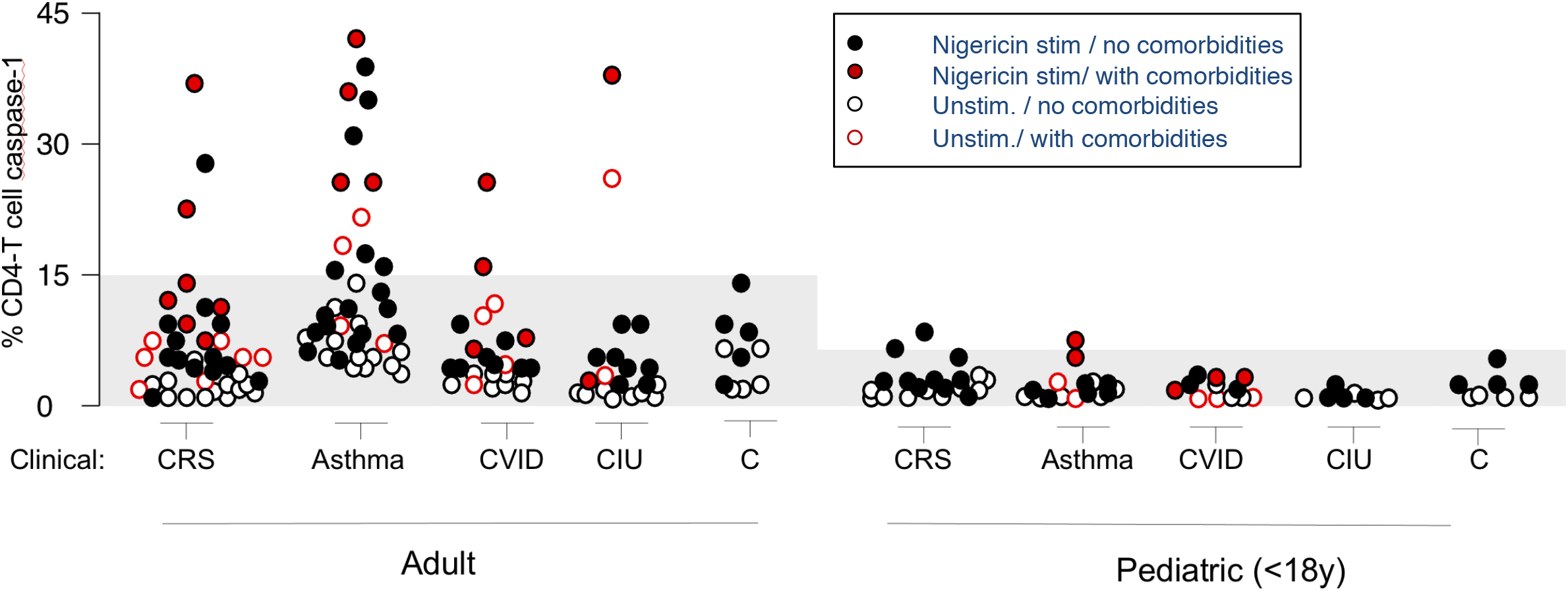
Caspase-1 expression on T cells in Non-COVID-19 patients (unexposed and uninfected adult and pediatric patients with allergic/immunological disorders). **Open circle** are resting non-stimulated CD4 T cells. **Closed circles** represent caspase 1 expression in nigericin stimulated CD4 T cells. **Red circles** represent patients with co-morbid conditions and (**Black circles)** represent patients without comorbidities. **Gray shaded** area represents the normal ranges for caspase-1+ CD4 T cell expression based on n=40 Healthy adults and n=40 <18years children.

### Caspase-1 up-regulation is not limited to the acute stage of COVID-19 disease

Up to 87% of inpatients and 35% of outpatients who recover from COVID-19 report persistence of at least 1 symptom, particularly fatigue and dyspnea (29, 30). Although preliminary reports describe this new feature as “post-COVID syndrome”, its mechanisms and natural history remains unknown. We assayed caspase-1 expression on the CD4 T cells of health care workers (HCWs) with persistent symptoms at least 90 days post-SARS-CoV-2 infection (Supplemental Table 4). There was significant up-regulation of baseline as well as nigericin stimulated T-helper cell caspase-1 levels only in symptomatic “post-COVID-19” HCWs, also known as long haulers (Figure 4). Interestingly, PCR-negative symptomatic HCWs with history of flu-like illness in early 2020 as well as those with positive IgG to SARS-CoV-2 also had increase caspase-1 expression. The level of expression of nigericin stimulated caspase-1 was comparable to those with active infection as seen in Figure 2, although the baseline caspase-1 levels were lower in these long haulers. Non-exposed control subjects showed no T cell caspase-1 overexpression.

**Figure 4.**
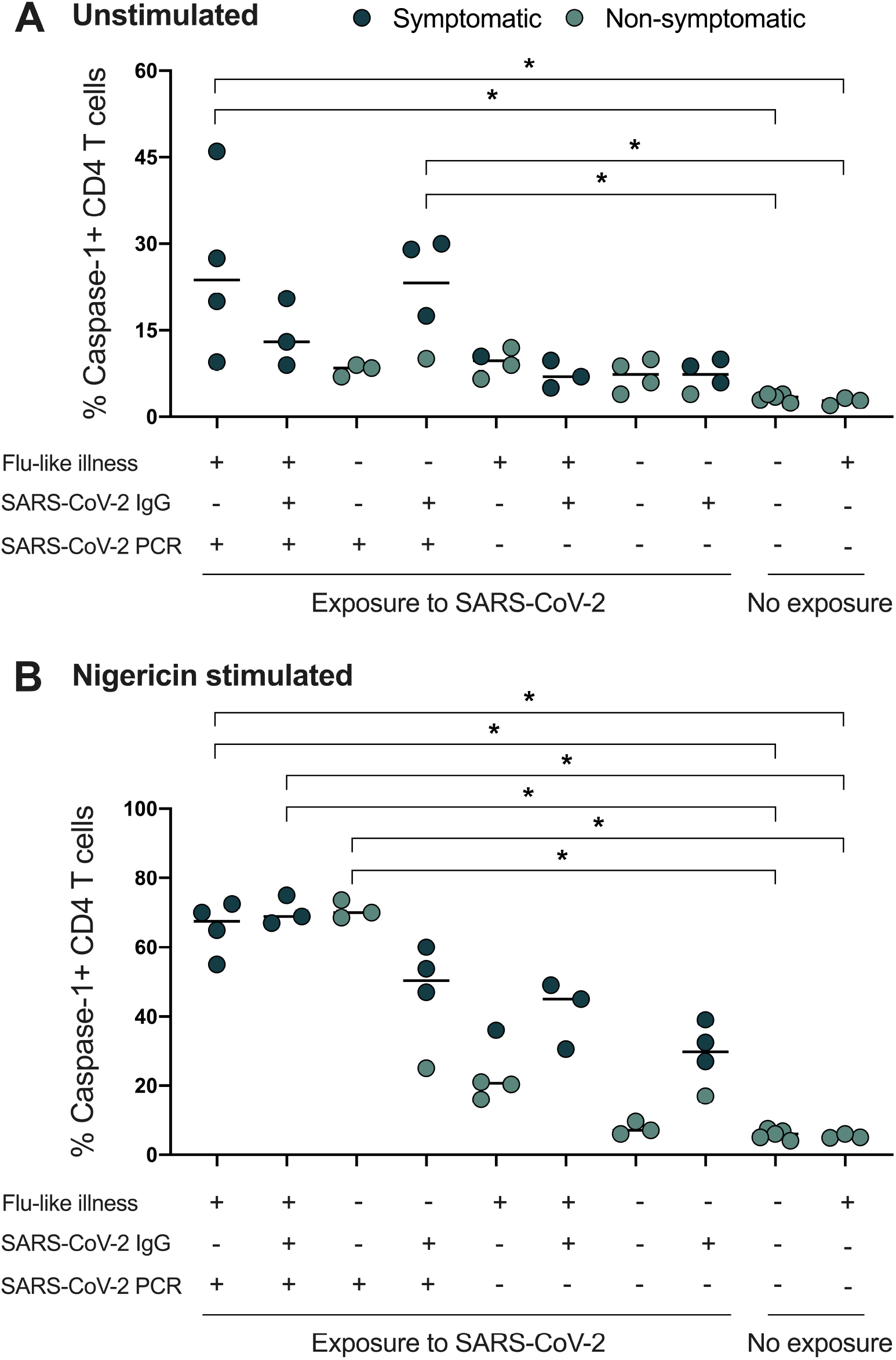
Caspase-1 expression on T cells in post-COVID-19 Health Care Workers. Blood samples were analyzed at least 90 days after SARS-CoV2 exposure in healthcare workers. Patients with no exposure history and negative PCR to SARS-CoV-2 were used as controls. **Solid circles** represent nigericin stimulated cells. **Red circles** represent symptomatic, black represent non-symptomatic patients. Exposure indicates being in close proximity to SARS-CoV-2 infected patients in the absence of personal protection equipment.

### Pan-caspase inhibitor suppresses elevated caspase-1 activity in CD4 T cells derived from moderate-severe COVID-19 patients

To assess whether CD4 T cell caspase-1 activity can be suppressed by small molecule caspase inhibitors, we incubated whole blood samples with either the oral pan-caspase inhibitor emricasan (EMR) (31) or the selective orally active ICE/caspase-1 inhibitor VX765 (32), followed 24hrs later with or without nigericin stimulation. We found that EMR suppressed CD4 T cell caspase-1 activity in COVID-19 samples or prevented its upregulation in healthy subjects (Figure 5), while VX765 did not show a suppressive effect.

**Figure 5.**
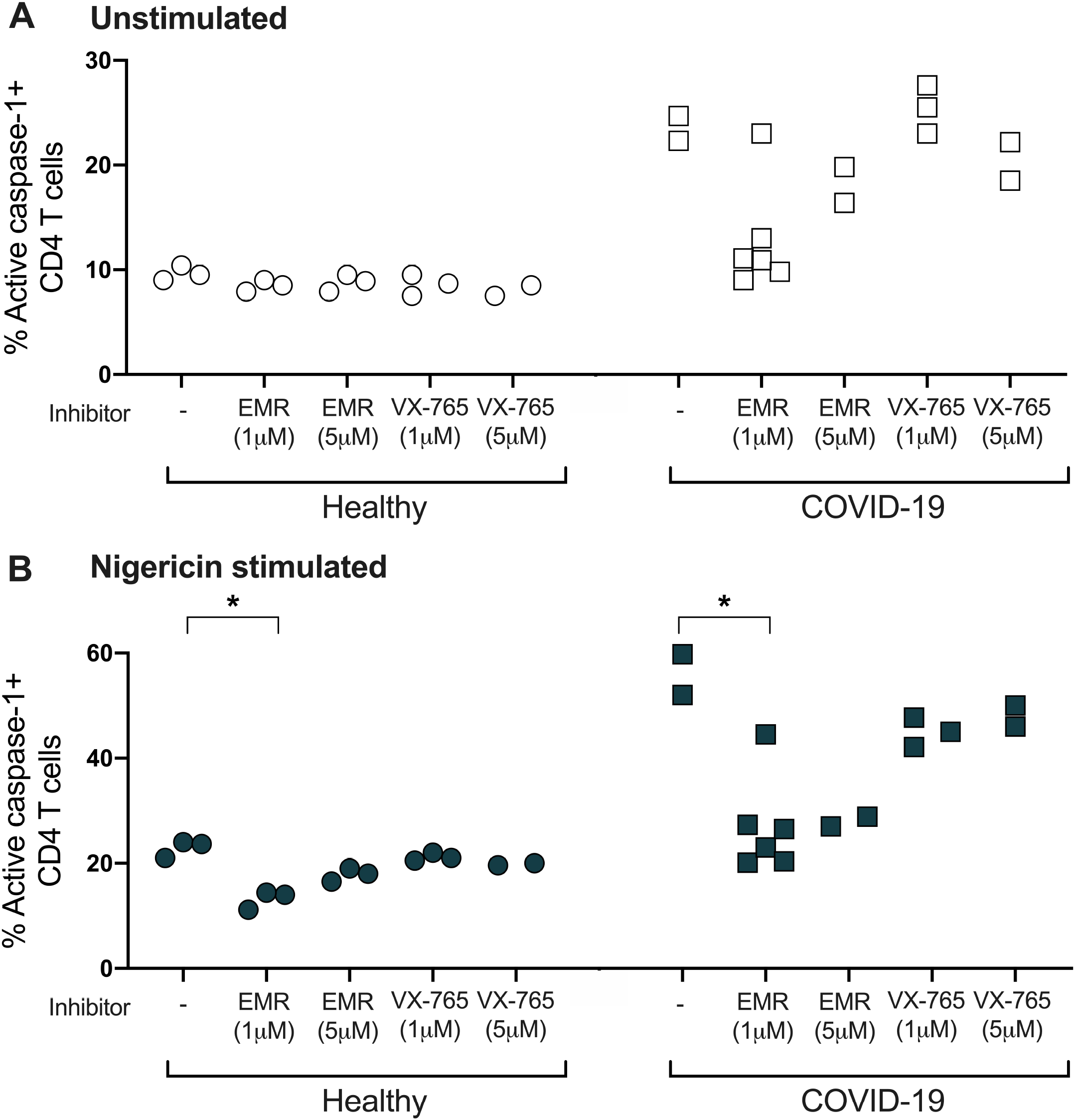
Effects of Caspase inhibitors on CD4 T cells in COVID-19 patients. Incubation of healthy and COVID-19 subjects samples with caspase inhibitors: EMR or VX765. Caspase-1 expression was measured by flow cytometry.

### Red Blood cells show increased caspase-3 in COVID-19 disease which is suppressed by a pan-caspase inhibitor

Recent reports suggest abnormalities in the RBCs in patients with COVID-19 (33-35). In the process of Ficoll separation, we observed a layer of RBCs contaminating the PBMC layer that was universally present in all samples from COVID-19 individuals (Figure 6A). This finding was also present in up to 80% of COVID-19 convalescent subjects. Plasma from acutely infected subjects induced a similar finding when incubated overnight with plasma depleted whole blood of healthy patients. Treatment of the plasma samples with trypsin, DNAse, or heat inactivation did not abolish this effect. Cellular caspases are not limited to immune cells. RBCs do not express caspase-1, but have been shown to have detectable caspase-3 that increases with various disorders (33). We found that RBCs from acute COVID-19 subjects showed significant up-regulation of caspase-3 compared to healthy controls (Figure 6B). Plasma from these patients also upregulated caspase-3 in healthy subjects’ RBCs. We did not see this effect when healthy subjects’ RBCs were incubated with plasma from influenza infected patients, although a similar RBC contamination was observed in these samples after Ficoll separation. Furthermore, EMR suppressed the caspase-3 up-regulation in the COVID-19 plasma incubated samples, but did not change the baseline expression levels in influenza-plasma incubated samples.

**Figure 6.**
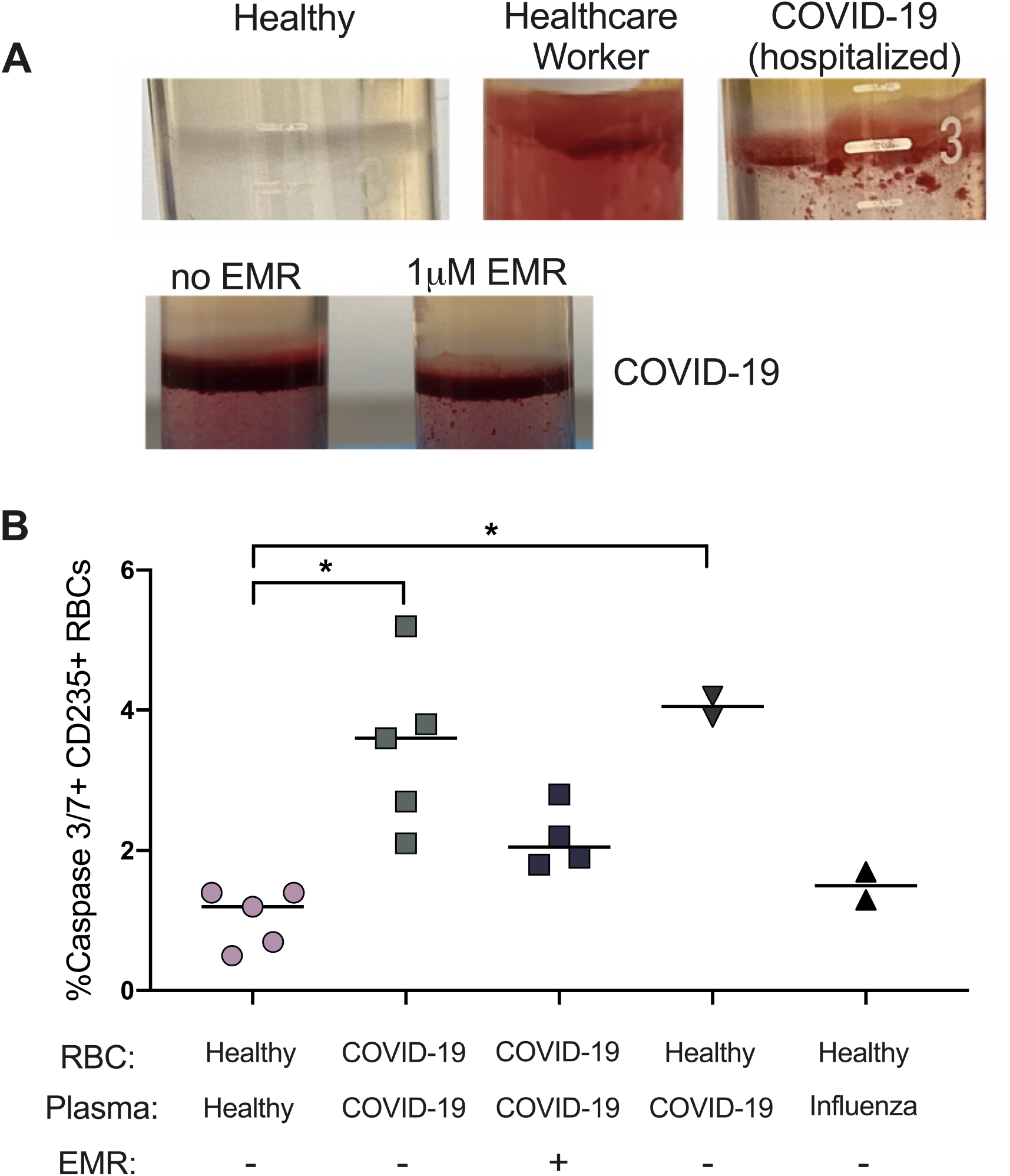
Caspase 3/7 expression in Red Blood cells derived from COVID-19 patients’. Blood samples were analyzed from hospitalized patients with SARS-CoV-2 infection. **A)** RBC contamination of the PBMC layer after Ficoll separation. **B)** Analysis of caspase 3/7 expression in COVID-19 patients and healthy controls. Some experiments were done using plasma from COVID-19 or subjects with influenza with incubated with RBCs from health uninfected donors as indicated.

## Discussion

While COVID-19 disease largely presents with respiratory symptoms, for many patients it is a systemic disease with a wide range of effects on different organ systems and persistent post-infection sequelae. A minority of children present with an autoimmune syndrome suggesting it is already a systemic disease for some people at the initial presentation (36-38). The blur between acute infections and post-infectious sequelae will only be understood better as detailed follow ups are performed along with immunological testing in natural history studies (39). In this report, we show preliminary evidence for caspase involvement that may shed further light not only into the systemic nature of the illness, but also the chronicity that is observed in COVID-19 long haulers.

There is accumulating evidence on the role of both apoptotic and pyroptotic cell death in the disease progression seen in COVID-19 (40). Pyroptosis leads to the production of IL-18 and IL-1β as a result of their cleavage by inflammasome activated caspase-1 (23, 41). IL-18 induces an IFN-*γ* response, while IL-1β induces neutrophil influx and activation, T and B-cell activation, cytokine and antibody production, and promotes Th17 differentiation (42-45). High levels of IL-18, IL-1β, and other proinflammatory cytokines were observed from the lungs and sera of COVID-19 patients (46). Although the ultimate outcome of activation of the inflammasome in the infected tissue cells (e.g. respiratory epithelial cells) should result in an enhanced immunity against pathogens, an accompanying pyroptosis and apoptosis of immune cells (e.g. T cell and macrophage/dendritic cells) also sending out danger signals may lead to poor outcomes overall. We hypothesized that T cell lymphopenia, which is a pathognomonic feature for SARS-CoV-2, might be a path in which the virus evades the human host response by creating an adaptive immune defect along with fueling an uncontrolled inflammatory response by the release of cellular contents of both immune and non-immune cells (47). Since caspase-1 is up-regulated predominantly in the memory CD4 T cell, this could explain why older persons are more susceptible to severe disease compared to the younger population. The end result is likely a self-damaging shut down of the immune system that further fuels the inflammation created by the viral infection, resulting in acute virus-induced immune deficiency (AVID). This connection between caspase-1 and COVID-19 has significant therapeutic implications since preventing the pyroptotic lymphocyte death rather than inhibition of the inflammatory response may be a more ideal option. The failure of cytokine targeted therapies could therefore suggest that adaptive immune dysfunction weighs more than an inflammatory response in disease progression (48).

A dysregulated caspase expression profile of immune cells can impact outcomes of an immune response. Caspases not only play an essential role during apoptotic cell death, but a subfamily of them, the inflammatory caspases, are associated with immune responses to microbial pathogens(49). Caspases can be subdivided into initiator and effector caspases. The subset of caspases that cleave selected substrates to produce the changes associated with apoptosis are known as effector caspases, which in mammals are caspase-3, -6, and -7. In most instances, these executioner caspases are activated by the initiator caspases, i.e., caspase-8, caspase-10, caspase-2, and caspase-9. Some of the “apoptotic” caspases, in particular caspase-8, have been shown to have additional roles in proliferation and differentiation. For example, caspase-8 activity appears to be necessary for lymphocyte proliferation (50). The clinical consequences of this dysregulated caspase activation can correlate with the hematological and immunological findings in COVID-19 patients (51). Although viral illnesses typically will impact the function or the life-cycle of lymphocytes, presenting with either lymphocytosis (for example CMV, influenza, varicella) or more rarely, lymphopenia (for example in H5N1, H1N1, HIV), the finding of neutrophilia in the setting of COVID-19 has been a common, but intriguing finding(52). The unique combination of inflammatory and apoptotic caspase upregulation coupled to the timing of the activation in the neutrophil response to SARS-CoV-2 may result in this unique laboratory finding observed in COVID-19.

An extensive body of evidence from the past three decades has implicated dysregulated caspase activation as a causal disease mechanism in tumorigenesis, autoimmunity, autoinflammation and infectious pathologies. As research into signaling pathways of inflammatory caspases progressed, an unexpected amount of crosstalk with apoptotic caspases has emerged over the past years. Pathogens use specific caspase pathways to evade the host, such as caspase-8 and RIPK1 for cell death and inflammasome responses induced by influenza A virus (53). Although caspases are most often associated with apoptosis, there has been persistent evidence that some of these enzymes can also influence proliferation (54). One of the earliest observations was that treatment of T cells with caspase inhibitors led to a surprising suppression of CD3-induced T-cell expansion. This growth-promoting caspase function was later attributed to caspase-8, because c-FLIP, a caspase-8 inhibitor, was shown to modulate T-cell proliferation. Similarly, caspase-8 and -6 can positively regulate B-cell proliferation (55). The suppression of caspase-8 in COVID-19 can be one mechanism through which the virus keeps the host immune responses in check. However, caspase-3 may have the opposite effect, as B cells lacking caspase-3 showed increased proliferation in vivo and hyperproliferation after mitogenic stimulation in vitro (56). Although the impact on caspase-3 is not seen in the immune cells circulating in the blood, tissue macrophages in postmortem analysis and circulating red blood cells do show significant increases. Caspase activity can be a double-edged sword and SARS CoV2 may be using this strategy to evade the host in way that’s beneficial for its own survival to move on to the next host to infect.

Little is known about the natural stimuli that lead to the assembly of complexes activating inflammatory caspases. The only physiological stimulus that is reported to activate caspase-1 in phagocytes is LPS (57). Very few studies extended this model of caspase-1 activation, mainly because it only induces moderate caspase-1 activity. The best-studied model of caspase-1 activation relates to the exposure of cells to extracellular ATP acting on P2×7 receptors. P2×7 receptors belong to a family of ion channel receptors gated by extracellular ATP and widely distributed in non-neuronal cells (57). Hence, at least two mechanisms are able to trigger activation of caspase-1, one as a consequence of a bacterial product and the other following changes in the intracellular ionic environment. It is of interest to see SARS-CoV-2 added to this list and further elucidating mechanisms on how this virus activates caspase-1 will be of importance in understanding disease mechanisms in COVID-19.

Taken together, our findings implicate that an induced acute immunodeficiency resulting from the necrotic cell death of lymphocytes may play an important factor in COVID-19 progression. We propose that the inflammatory response is secondary to the “danger signals” from necrotic cell death of immune system cells, resulting in a heightened inflammation compared to the one induced by dying tissue cells (58-60). The end result is likely a self-damaging cell death and subsequent cytokine storm that further fuels the inflammation created by the viral infection, resulting in AVID. The failure of cytokine targeted therapies could be due to that adaptive immune dysfunction weighs more heavily than an inflammatory response in disease progression (15). This connection between caspase-1 and COVID-19 has significant therapeutic implications since preventing the pyroptotic lymphocyte death rather than inhibition of the inflammatory response may be a more efficacious therapeutic option. Furthermore, an optimal therapeutic approach would be to also prevent progression into this end-stage disease by the use of therapeutics targeting the virus, pyroptosis or both.

The caspase mediated disease process in COVID-19 is most likely not limited to caspase-1 and T cells, as we demonstrate changes in RBCs that may additionally involve the caspase-3 pathway. Inflammatory microvascular thrombi are present in the lung, kidney, and heart, containing neutrophil extracellular traps associated with platelets and fibrin along with changes in RBC morphology (61, 62), which raise the possibility of RBCs playing a role in the clotting disorder observed in COVID-19 patients. Although the RBC layer contaminating the PBMCs in COVID-19 blood samples is also observed in other infections, such as influenza, the caspase-3 upregulation may lead to changes in the RBC that may mechanistically link them to complications of the disease (63-65).

Both metabolic inflammation and apoptosis play a central role in the pathogenesis of metabolic disease such as obesity, diabetes and the progression of nonalcoholic steatohepatisis (NASH) to more severe liver disease (66-68). Caspase-1-dependent inflammasome activation has been shown to have a crucial function in the establishment of diabetic nephropathy (69). In an animal model of hypertension apoptosis of myocardial cells were demonstrated, and the apoptosis becomes more serious with the constantly elevated level and prolonged duration of hypertension. The activity of caspase-3 was shown to have a close correlation with cardiomyocyte apoptosis (70). The pan-caspase inhibitor, EMR has been shown in a bioinformatics computational screen to binding to the COVID-19 receptor ACE2, suggesting the potential to block cell entry (71). In a separate unrelated study, a screen of ∼6,070 drugs with a known 28 previous history of use in humans was conducted to identify compounds that inhibit the activity of SARS-CoV-2 main protease Mpro *in vitro* (72). EMR was shown to be among 50 compounds with activity against Mpro with an overall hit rate <0.75%. As an oral formulation, EMR has been shown to be well-tolerated, reduced serum markers of apoptosis (caspase-3/7), liver enzymes, function (e.g. reducing ALT, MELD & Child-Pugh scores, INR and total bilirubin) and inflammatory biomarkers (CK-18) in patients w/ hepatitis C virus and NASH. There was no impact on histology of the disease characterized by liver fibrosis. (73). Lack of improvement in liver histology with EMR, despite such a dramatic and universal reduction in serum biomarkers of disease activity suggest a mechanisms that lead to liver injury, resulting in increase in liver enzymes that EMR has a positive impact on is different than those causing fibrosis in NASH (74, 75). One can also consider an overwhelming cellular injury that EMR is not able to reverse, or lack of efficacy due to therapy being initiated late in the course of the liver disease.

Our findings support a novel alternate therapeutic approach against COVID-19 through the use of a caspase inhibitor early on in the course of infection to alleviate or prevent disease progression. Although SARS-CoV-2 does not seem to infect immune system cells (with the possible exception of macrophage or dendritic cells), the outcome of T cell depletion in severe forms of the disease seems to be through a similar mechanism to that seen in HIV; caspase-1 activation (76). Perhaps it will be important to better understand the impact of different co-morbid conditions on T cell caspase expression at baseline, before exposure to SARS-CoV-2, which may be the determining factor for developing severe disease. There is a large body of evidence pointing out to an activated inflammasome in a wide variety of disorders that overlap with high-risk conditions for severe COVID-19 (13, 59, 60, 77). Ultimately *in vivo* clinical data is necessary to test the hypothesis of whether pan-caspase inhibition can prevent inflammasome activation in early onset SARS-CoV-2 patients and subsequent lymphopenia and sequelae development. Preliminary evidence on the therapeutic effect of EMR on SARS-CoV-2 viral protease and ACE2 receptor inhibition raise a relevant key question that will need to be answered through a randomized clinical trial on the multi-modal actions of this pan-caspase inhibitor in the setting of COVID-19.

## Data Availability

All data regarding the manuscript is available on request

## Supplemental Figures and Tables

**Supplemental Figure 1. RNA-Seq data from cell lines infected *in vitro* with SARS-CoV-2**. Expression values for Caspase 1, 4, and 5 genes were normalized by DESeq2

**Supplemental Figure 2. RNA-Seq data from cell lines infected *in vitro* with SARS-CoV-2**. Expression values for Caspase 2, 8 amd 9 genes were normalized by DESeq2

**Supplemental Figure 3. RNA-Seq data from cell lines infected *in vitro* with SARS-CoV-2**. Expression values for Caspase 3, 6 and 7 genes were normalized by DESeq2

**Supplemental Table 1. Co-morbidities of Critical/ICU and non-Critical/non-ICU COVID-19 patients**.

**Supplemental Table 2. Lymphocyte immune phenotyping results of healthy and COVID-19 subjects and correlation with CD3+CD4+**, **CD3+ and CD3-caspase+ T cells**.

**Table 1.** Demographics of hospitalized (Critical/ICU and non-Critical/non-ICU) COVID-19 patients and healthy subject

|  | **HOSPITALIZED** | *SARS-CoV2 PCR +* | **HEALTHY** |
| --- | --- | --- | --- |
|  | **Critical/ICU** | **non-critical/Non ICU** | *SARS CoV2 Ab*<br>*Neg* |
| n | 14 | 11 | 14 |
| Age (median) | 64 | 71.5 | 54 |
| 80+ | 3 | 1 | 3 |
| 71-80 | 3 | 3 | 2 |
| 61-70 | 1 | 2 | 4 |
| 51-60 | 3 | 3 | 1 |
| 41-50 | 2 | 1 | 3 |
| 0-40 | 2 | 1 | 1 |
| Ethnicity |  |  |  |
| african/american | 7 | 4 | 4 |
| other | 7 | 7 | 10 |
| Gender |  |  |  |
| male | 6 | 5 | 7 |
| female | 8 | 6 | 7 |
| mean BMI | 31 | 29 | 26 |

**Table 2.**
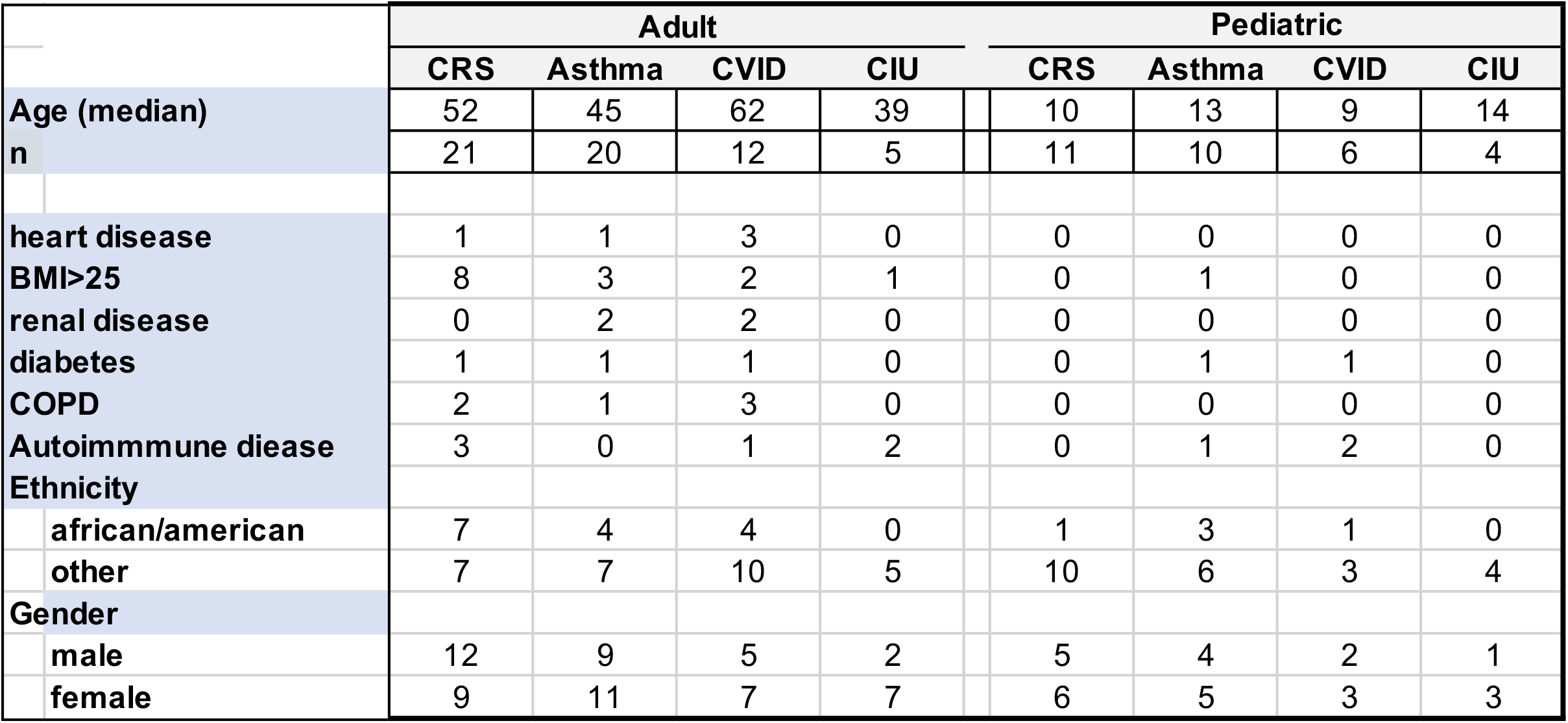
Demographics of Adult and Pediatric non-COVID-19 patient

| Table 2 |  |  |  |  |  |  |  |  |  |
| --- | --- | --- | --- | --- | --- | --- | --- | --- | --- |
|  |  | Adult |  |  |  | Pediatric |  |  |  |
|  |  | CRS | Asthma | CVID | CIU | CRS | Asthma | CVID | CIU |
| Age (median) |  | 52 | 45 | 62 | 39 | 10 | 13 | 9 | 14 |
| n |  | 21 | 20 | 12 | 5 | 11 | 10 | 6 | 4 |
| heart disease |  | 1 | 1 | 3 | 0 | 0 | 0 | 0 | 0 |
| BMI>25 |  | 8 | 3 | 2 | 1 | 0 | 1 | 0 | 0 |
| renal disease |  | 0 | 2 | 2 | 0 | 0 | 0 | 0 | 0 |
| diabetes |  | 1 | 1 | 1 | 0 | 0 | 1 | 1 | 0 |
| COPD |  | 2 | 1 | 3 | 0 | 0 | 0 | 0 | 0 |
| Autoimmune disease |  | 3 | 0 | 1 | 2 | 0 | 1 | 2 | 0 |
| Ethnicity |  |  |  |  |  |  |  |  |  |
| african/american |  | 7 | 4 | 4 | 0 | 1 | 3 | 1 | 0 |
| other |  | 7 | 7 | 10 | 5 | 10 | 6 | 3 | 4 |
| Gender |  |  |  |  |  |  |  |  |  |
| male |  | 12 | 9 | 5 | 2 | 5 | 4 | 2 | 1 |
| female |  | 9 | 11 | 7 | 7 | 6 | 5 | 3 | 3 |

**Table 3.** Demographics of Healthcare workers

|  | **HCW** | **Co-morbidities** |
| --- | --- | --- |
| **n** | 36 |  |
| **Age (median)** | 64 |  |
| 71-80 | 3 (8) | 2 (67) |
| 61-70 | 4 (11) | 2 (50) |
| 51-60 | 16 (44) | 8 (50) |
| 41-50 | 4 (11) | 1 (25) |
| 18-40 | 9 (25) | 2 (22) |
| **Ethnicity** |  |  |
| african/american | 12 (33) | 7 (58) |
| other | 24 (67) | 8 (33) |
| **Gender** |  |  |
| male | 15 (42) | 9 (60) |
| female | 21 (68) | 6 (40) |

## Notes

### Competing Interest Statement

None of the authors have any relevant conflict of interest to disclose with the exception of L.C.N. has received fees from Abbvie within the past year and has received compensation for serving on a scientific advisory board for Cytodyn for work unrelated to this project. All other authors declare no competing interests.

### Funding Statement

None for the study. L.C.N. is supported by several NIH awards including from the National Institute of Mental Health (R01 MH112457) and National Institute of Allergy and Infectious Diseases (UM1 AI126617) BELIEVE Collaboratories.

### Author Declarations

Western IRB: 1285028 for Amerimmune S.U.N.Y. Downstate IRB 269846-10 for SUNY Downstate Medical Center

## References

1. Fauci AS, Lane HC, and Redfield RR. Covid-19 - Navigating the Uncharted. N Engl J Med. 2020;382(13):1268–9.

2. Paules CI, Marston HD, and Fauci AS. Coronavirus Infections-More Than Just the Common Cold. JAMA. 2020;323(8):707–8.

3. Barlow A, Landolf KM, Barlow B, Yeung SYA, Heavner JJ, Claassen CW, et al. Review of Emerging Pharmacotherapy for the Treatment of Coronavirus Disease 2019. Pharmacotherapy. 2020;40(5):416–37.

4. Xu X, Han M, Li T, Sun W, Wang D, Fu B, et al. Effective treatment of severe COVID-19 patients with tocilizumab. Proc Natl Acad Sci U S A. 2020;117(20):10970–5.

5. Spinner CD, Gottlieb RL, Criner GJ, Arribas Lopez JR, Cattelan AM, Soriano Viladomiu A, et al. Effect of Remdesivir vs Standard Care on Clinical Status at 11 Days in Patients With Moderate COVID-19: A Randomized Clinical Trial. JAMA. 2020;324(11):1048–57.

6. Alijotas-Reig J, Esteve-Valverde E, Belizna C, Selva-O’Callaghan A, Pardos-Gea J, Quintana A, et al. Immunomodulatory therapy for the management of severe COVID-19. Beyond the anti-viral therapy: A comprehensive review. Autoimmun Rev. 2020;19(7):102569.

7. Baum A, Fulton BO, Wloga E, Copin R, Pascal KE, Russo V, et al. Antibody cocktail to SARS-CoV-2 spike protein prevents rapid mutational escape seen with individual antibodies. Science. 2020;369(6506):1014–8.

8. Soy M, Keser G, Atagunduz P, Tabak F, Atagunduz I, and Kayhan S. Cytokine storm in COVID-19: pathogenesis and overview of anti-inflammatory agents used in treatment. Clin Rheumatol. 2020;39(7):2085–94.

9. Becker RC. COVID-19 update: Covid-19-associated coagulopathy. J Thromb Thrombolysis. 2020;50(1):54–67.

10. Hasoksuz M, Kilic S, and Sarac F. Coronaviruses and SARS-COV-2. Turk J Med Sci. 2020;50(SI-1):549–56.

11. Pouletty M, Borocco C, Ouldali N, Caseris M, Basmaci R, Lachaume N, et al. Paediatric multisystem inflammatory syndrome temporally associated with SARS-CoV-2 mimicking Kawasaki disease (Kawa-COVID-19): a multicentre cohort. Ann Rheum Dis. 2020;79(8):999–1006.

12. Wu JT, Leung K, Bushman M, Kishore N, Niehus R, de Salazar PM, et al. Estimating clinical severity of COVID-19 from the transmission dynamics in Wuhan, China. Nat Med. 2020;26(4):506–10.

13. Huang C, Wang Y, Li X, Ren L, Zhao J, Hu Y, et al. Clinical features of patients infected with 2019 novel coronavirus in Wuhan, China. Lancet. 2020;395(10223):497–506.

14. Mehta P, McAuley DF, Brown M, Sanchez E, Tattersall RS, Manson JJ, et al. COVID-19: consider cytokine storm syndromes and immunosuppression. Lancet. 2020;395(10229):1033–4.

15. Kroemer A, Khan K, Plassmeyer M, Alpan O, Haseeb MA, Gupta R, et al. Inflammasome activation and pyroptosis in lymphopenic liver patients with COVID-19. J Hepatol. 2020.

16. Aid M, Busman-Sahay K, Vidal SJ, Maliga Z, Bondoc S, Starke C, et al. Vascular Disease and Thrombosis in SARS-CoV-2-Infected Rhesus Macaques. Cell. 2020.

17. Gao YL, Zhai JH, and Chai YF. Recent Advances in the Molecular Mechanisms Underlying Pyroptosis in Sepsis. Mediators Inflamm. 2018;2018:5823823.

18. Jia C, Chen H, Zhang J, Zhou K, Zhuge Y, Niu C, et al. Role of pyroptosis in cardiovascular diseases. Int Immunopharmacol. 2019;67:311–8.

19. Man SM, Karki R, and Kanneganti TD. Molecular mechanisms and functions of pyroptosis, inflammatory caspases and inflammasomes in infectious diseases. Immunol Rev. 2017;277(1):61–75.

20. Kovacs SB, and Miao EA. Gasdermins: Effectors of Pyroptosis. Trends Cell Biol. 2017;27(9):673–84.

21. Zhang Y, Chen X, Gueydan C, and Han J. Plasma membrane changes during programmed cell deaths. Cell Res. 2018;28(1):9–21.

22. Fleisher TA. Apoptosis. Ann Allergy Asthma Immunol. 1997;78(3):245-9; quiz 9-50.

23. Moretti J, and Blander JM. Increasing complexity of NLRP3 inflammasome regulation. J Leukoc Biol. 2020.

24. Jamilloux Y, Henry T, Belot A, Viel S, Fauter M, El Jammal T, et al. Should we stimulate or suppress immune responses in COVID-19? Cytokine and anti-cytokine interventions. Autoimmun Rev. 2020;19(7):102567.

25. Al-Samkari H, Karp Leaf RS, Dzik WH, Carlson JCT, Fogerty AE, Waheed A, et al. COVID-19 and coagulation: bleeding and thrombotic manifestations of SARS-CoV-2 infection. Blood. 2020;136(4):489–500.

26. Grobler C, Maphumulo SC, Grobbelaar LM, Bredenkamp JC, Laubscher GJ, Lourens PJ, et al. Covid-19: The Rollercoaster of Fibrin(Ogen), D-Dimer, Von Willebrand Factor, P-Selectin and Their Interactions with Endothelial Cells, Platelets and Erythrocytes. Int J Mol Sci. 2020;21(14).

27. Wilk AJ, Rustagi A, Zhao NQ, Roque J, Martinez-Colon GJ, McKechnie JL, et al. A single-cell atlas of the peripheral immune response in patients with severe COVID-19. Nat Med. 2020;26(7):1070–6.

28. Blanco-Melo D, Nilsson-Payant BE, Liu WC, Uhl S, Hoagland D, Moller R, et al. Imbalanced Host Response to SARS-CoV-2 Drives Development of COVID-19. Cell. 2020;181(5):1036–45 e9.

29. Carfi A, Bernabei R, Landi F, and Gemelli Against C-P-ACSG. Persistent Symptoms in Patients After Acute COVID-19. JAMA. 2020;324(6):603–5.

30. Garg P, Arora U, Kumar A, and Wig N. The “post-COVID” syndrome: How deep is the damage? J Med Virol. 2020.

31. Linton SD, Aja T, Armstrong RA, Bai X, Chen LS, Chen N, et al. First-in-class pan caspase inhibitor developed for the treatment of liver disease. J Med Chem. 2005;48(22):6779–82.

32. Stack JH, Beaumont K, Larsen PD, Straley KS, Henkel GW, Randle JC, et al. IL-converting enzyme/caspase-1 inhibitor VX-765 blocks the hypersensitive response to an inflammatory stimulus in monocytes from familial cold autoinflammatory syndrome patients. J Immunol. 2005;175(4):2630–4.

33. Foy BH, Carlson JCT, Reinertsen E, Padros IVR, Pallares Lopez R, Palanques-Tost E, et al. Association of Red Blood Cell Distribution Width With Mortality Risk in Hospitalized Adults With SARS-CoV-2 Infection. JAMA Netw Open. 2020;3(9):e2022058.

34. Maellaro E, Leoncini S, Moretti D, Del Bello B, Tanganelli I, De Felice C, et al. Erythrocyte caspase-3 activation and oxidative imbalance in erythrocytes and in plasma of type 2 diabetic patients. Acta Diabetol. 2013;50(4):489–95.

35. Thomas T, Stefanoni D, Dzieciatkowska M, Issaian A, Nemkov T, Hill RC, et al. Evidence for structural protein damage and membrane lipid remodeling in red blood cells from COVID-19 patients. medRxiv. 2020.

36. Cheung EW, Zachariah P, Gorelik M, Boneparth A, Kernie SG, Orange JS, et al. Multisystem Inflammatory Syndrome Related to COVID-19 in Previously Healthy Children and Adolescents in New York City. JAMA. 2020;324(3):294–6.

37. Jiang L, Tang K, Levin M, Irfan O, Morris SK, Wilson K, et al. COVID-19 and multisystem inflammatory syndrome in children and adolescents. Lancet Infect Dis. 2020;20(11):e276–e88.

38. Singh-Grewal D, Lucas R, McCarthy K, Cheng AC, Wood N, Ostring G, et al. Update on the COVID-19-associated inflammatory syndrome in children and adolescents; paediatric inflammatory multisystem syndrome-temporally associated with SARS-CoV-2. J Paediatr Child Health. 2020;56(8):1173–7.

39. Richardson S, Hirsch JS, Narasimhan M, Crawford JM, McGinn T, Davidson KW, et al. Presenting Characteristics, Comorbidities, and Outcomes Among 5700 Patients Hospitalized With COVID-19 in the New York City Area. JAMA. 2020;323(20):2052–9.

40. Yap JKY, Moriyama M, and Iwasaki A. Inflammasomes and Pyroptosis as Therapeutic Targets for COVID-19. J Immunol. 2020;205(2):307–12.

41. Sutterwala FS, Haasken S, and Cassel SL. Mechanism of NLRP3 inflammasome activation. Ann N Y Acad Sci. 2014;1319:82–95.

42. Akdis M, Aab A, Altunbulakli C, Azkur K, Costa RA, Crameri R, et al. Interleukins (from IL-1 to IL-38), interferons, transforming growth factor beta, and TNF-alpha: Receptors, functions, and roles in diseases. J Allergy Clin Immunol. 2016;138(4):984–1010.

43. Arend WP, Palmer G, and Gabay C. IL-1, IL-18, and IL-33 families of cytokines. Immunol Rev. 2008;223:20–38.

44. He Y, Hara H, and Nunez G. Mechanism and Regulation of NLRP3 Inflammasome Activation. Trends Biochem Sci. 2016;41(12):1012–21.

45. Latz E, Xiao TS, and Stutz A. Activation and regulation of the inflammasomes. Nat Rev Immunol. 2013;13(6):397–411.

46. Zhou Z, Ren L, Zhang L, Zhong J, Xiao Y, Jia Z, et al. Heightened Innate Immune Responses in the Respiratory Tract of COVID-19 Patients. Cell Host Microbe. 2020;27(6):883–90 e2.

47. Thompson E, Cascino K, Ordonez A, Zhou W, Vaghasia A, Hamacher-Brady A, et al. Mitochondrial induced T cell apoptosis and aberrant myeloid metabolic programs define distinct immune cell subsets during acute and recovered SARS-CoV-2 infection. medRxiv. 2020.

48. Buszko M, Park JH, Verthelyi D, Sen R, Young HA, and Rosenberg AS. The dynamic changes in cytokine responses in COVID-19: a snapshot of the current state of knowledge. Nat Immunol. 2020;21(10):1146–51.

49. Martinon F, and Tschopp J. Inflammatory caspases: linking an intracellular innate immune system to autoinflammatory diseases. Cell. 2004;117(5):561–74.

50. Feng Y, Daley-Bauer LP, and Mocarski ES. Caspase-8-dependent control of NK- and T cell responses during cytomegalovirus infection. Med Microbiol Immunol. 2019;208(3-4):555-71.

51. Terpos E, Ntanasis-Stathopoulos I, Elalamy I, Kastritis E, Sergentanis TN, Politou M, et al. Hematological findings and complications of COVID-19. Am J Hematol. 2020;95(7):834–47.

52. Tomar B, Anders HJ, Desai J, and Mulay SR. Neutrophils and Neutrophil Extracellular Traps Drive Necroinflammation in COVID-19. Cells. 2020;9(6).

53. Atkin-Smith GK, Duan M, Chen W, and Poon IKH. The induction and consequences of Influenza A virus-induced cell death. Cell Death Dis. 2018;9(10):1002.

54. McIlwain DR, Berger T, and Mak TW. Caspase functions in cell death and disease. Cold Spring Harb Perspect Biol. 2013;5(4):a008656.

55. Hakem A, El Ghamrasni S, Maire G, Lemmers B, Karaskova J, Jurisicova A, et al. Caspase-8 is essential for maintaining chromosomal stability and suppressing B-cell lymphomagenesis. Blood. 2012;119(15):3495–502.

56. Woo M, Hakem R, Furlonger C, Hakem A, Duncan GS, Sasaki T, et al. Caspase-3 regulates cell cycle in B cells: a consequence of substrate specificity. Nat Immunol. 2003;4(10):1016–22.

57. Franchi L, Eigenbrod T, Munoz-Planillo R, and Nunez G. The inflammasome: a caspase-1-activation platform that regulates immune responses and disease pathogenesis. Nat Immunol. 2009;10(3):241–7.

58. Labzin LI, Lauterbach MA, and Latz E. Interferons and inflammasomes: Cooperation and counterregulation in disease. J Allergy Clin Immunol. 2016;138(1):37–46.

59. Matzinger P. Tolerance, danger, and the extended family. Annu Rev Immunol. 1994;12:991–1045.

60. Matzinger P. The danger model: a renewed sense of self. Science. 2002;296(5566):301–5.

61. Zini G, Bellesi S, Ramundo F, and d’Onofrio G. Morphological anomalies of circulating blood cells in COVID-19. Am J Hematol. 2020;95(7):870–2.

62. Liu Y, Zhang X, Qiao J, Gong R, You Q, Sun J, et al. A Controllable Inflammatory Response and Temporary Abnormal Coagulation in Moderate Disease of COVID-19 in Wuhan, China. J Clin Med Res. 2020;12(9):590–7.

63. Carelli-Alinovi C, Pirolli D, Giardina B, and Misiti F. Protein kinase C mediates caspase 3 activation: A role for erythrocyte morphology changes. Clin Hemorheol Microcirc. 2015;59(4):345–54.

64. Firat U, Kaya S, Cim A, Buyukbayram H, Gokalp O, Dal MS, et al. Increased caspase-3 immunoreactivity of erythrocytes in STZ diabetic rats. Exp Diabetes Res. 2012;2012:316384.

65. Rinalducci S, Ferru E, Blasi B, Turrini F, and Zolla L. Oxidative stress and caspase-mediated fragmentation of cytoplasmic domain of erythrocyte band 3 during blood storage. Blood Transfus. 2012;10 Suppl 2:s55–62.

66. Wiewiora M, Piecuch J, Sedek L, Mazur B, and Sosada K. The effects of obesity on CD47 expression in erythrocytes. Cytometry B Clin Cytom. 2017;92(6):485–91.

67. Marini JJ, and Gattinoni L. Management of COVID-19 Respiratory Distress. JAMA. 2020;323(22):2329–30.

68. Neupane K, Ahmed Z, Pervez H, Ashraf R, and Majeed A. Potential Treatment Options for COVID-19: A Comprehensive Review of Global Pharmacological Development Efforts. Cureus. 2020;12(6):e8845.

69. Shahzad K, Bock F, Al-Dabet MM, Gadi I, Kohli S, Nazir S, et al. Caspase-1, but Not Caspase-3, Promotes Diabetic Nephropathy. J Am Soc Nephrol. 2016;27(8):2270–5.

70. Communal C, Sumandea M, de Tombe P, Narula J, Solaro RJ, and Hajjar RJ. Functional consequences of caspase activation in cardiac myocytes. Proc Natl Acad Sci U S A. 2002;99(9):6252–6.

71. Kim J, Zhang J, Cha Y, Kolitz S, Funt J, Escalante Chong R, et al. Advanced bioinformatics rapidly identifies existing therapeutics for patients with coronavirus disease-2019 (COVID-19). J Transl Med. 2020;18(1):257.

72. Jeremy D. Baker RLU, Gerald C. Kraemer, Jason E. Love, and Brian C. Kraemer. A drug repurposing screen identifies hepatitis C antivirals as inhibitors of the SARS-CoV-2 main 1 protease. 2020.

73. Frenette CT, Morelli G, Shiffman ML, Frederick RT, Rubin RA, Fallon MB, et al. Emricasan Improves Liver Function in Patients With Cirrhosis and High Model for End-Stage Liver Disease Scores Compared With Placebo. Clin Gastroenterol Hepatol. 2019;17(4):774–83 e4.

74. Barreyro FJ, Holod S, Finocchietto PV, Camino AM, Aquino JB, Avagnina A, et al. The pan-caspase inhibitor Emricasan (IDN-6556) decreases liver injury and fibrosis in a murine model of non-alcoholic steatohepatitis. Liver Int. 2015;35(3):953–66.

75. Harrison SA, Goodman Z, Jabbar A, Vemulapalli R, Younes ZH, Freilich B, et al. A randomized, placebo-controlled trial of emricasan in patients with NASH and F1-F3 fibrosis. J Hepatol. 2020;72(5):816–27.

76. Doitsh G, Galloway NL, Geng X, Yang Z, Monroe KM, Zepeda O, et al. Cell death by pyroptosis drives CD4 T-cell depletion in HIV-1 infection. Nature. 2014;505(7484):509–14.

77. Schurink B, Roos E, Radonic T, Barbe E, Bouman CSC, de Boer HH, et al. Viral presence and immunopathology in patients with lethal COVID-19: a prospective autopsy cohort study. Lancet Microbe. 2020.

