## supplemental data for "Caspases in COVID-19 Disease and Sequela and the Therapeutic Potential of Caspase Inhibitors"

**Supplemental Figure 1**

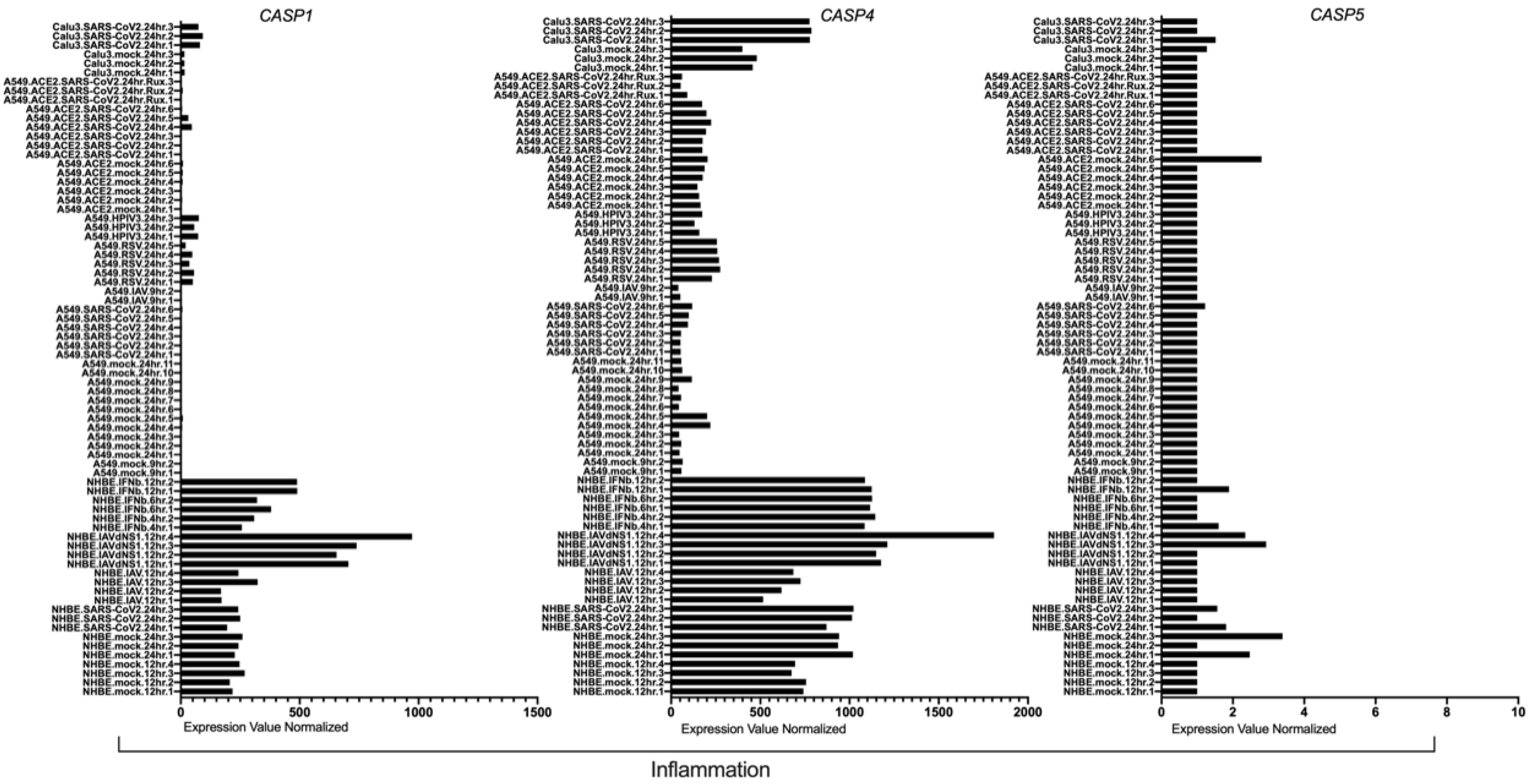

**Supplemental Figure 2**

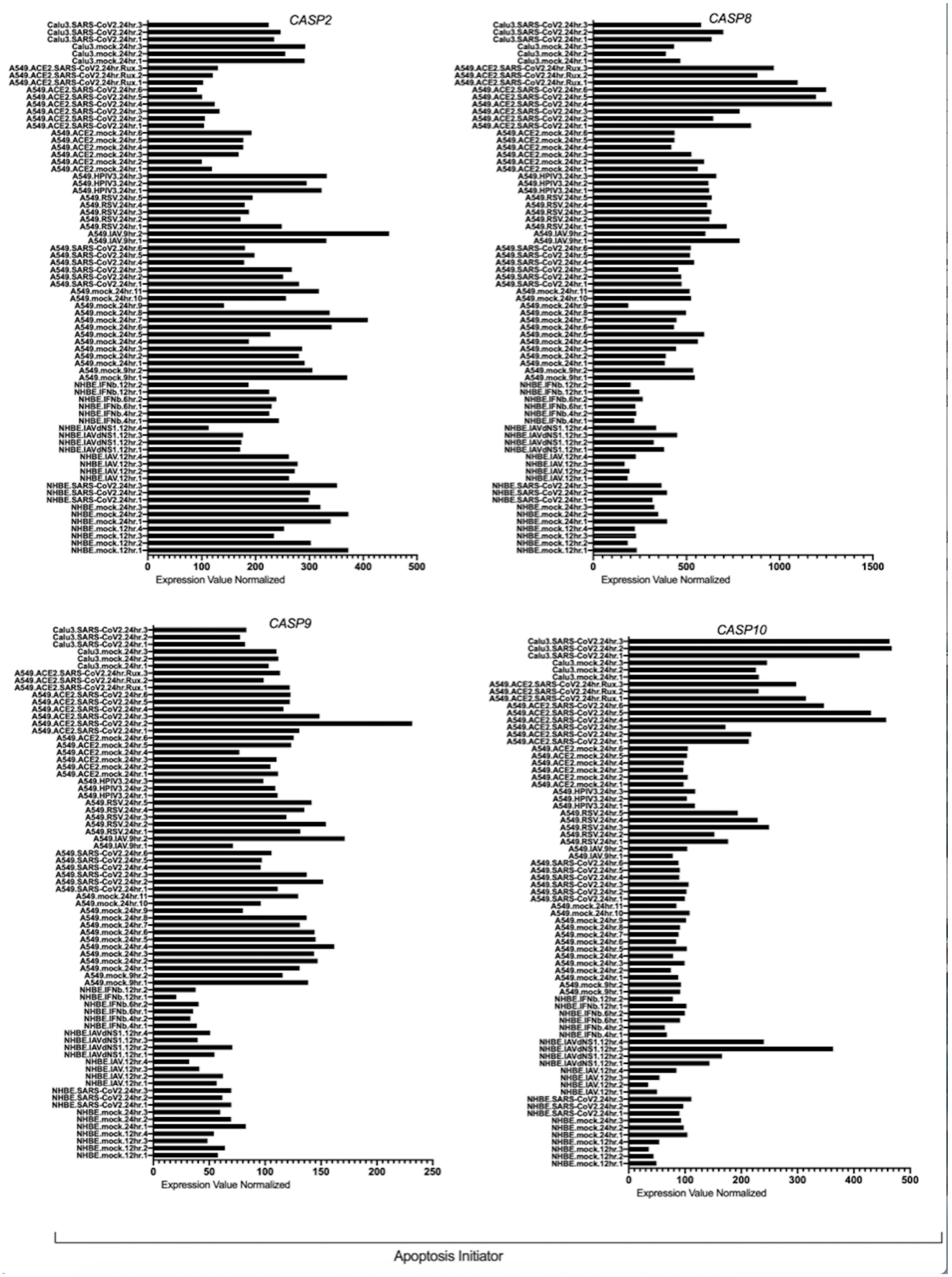

### Supplemental Figure 3

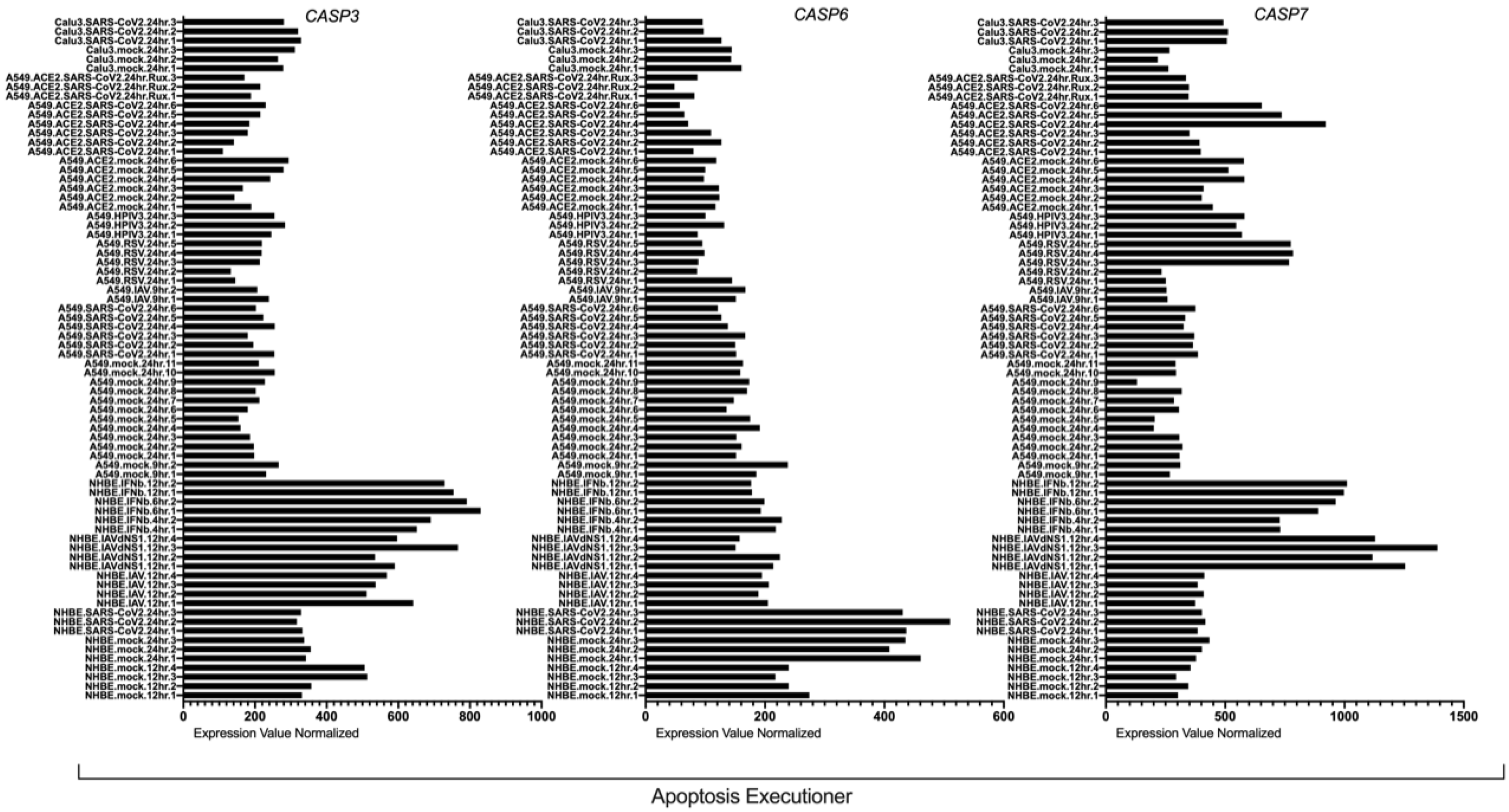

**Supp Table 1**

| | COVID-19 | |
|---|---|---|
| | ICU | Non-ICU |
| **Asthma** | 3 (21) | 3 (27) |
| **Autoimmune disease** | 3 (21) | 2 (18) |
| **Cancer** | 1 (7) | 3 (27) |
| **COPD** | 4 (29) | 3 (27) |
| **Coronary Artery Disease** | 2 (14) | 6 (55) |
| **Congestive Heart Failure** | 3 (21) | 5 (45) |
| **CKD without dialysis** | 2 (14) | 4 (36) |
| **CKD/ESRD with dialysis** | 3 (21) | 2 (18) |
| **Diabetes Mellitus** | 4 (29) | 4 (36) |
| **Hyperlipidemia** | 4 (29) | 5 (45) |
| **Hypertension** | 6 (43) | 5 (45) |
| **Immune suppression** | 5 (36) | 4 (36) |

#### Supplemental Table 2.

| Lymphocyte Monitoring | 18+ Normal Range | Healthy Mean | Healthy SEM | Covid-19 + Mean | Covid-19 + SEM | Covid-19+ ICU Mean | Covid-19+ ICU SEM | Covid-19+ Non ICU Mean | Covid-19+ Non ICU SEM |
|---|---|---|---|---|---|---|---|---|---|
| Monocytes | 3.1 - 11.3 | 5.86 | 0.30 | 5.78 | 0.45 | 4.06 | 0.37 | 7.82 | 0.62 |
| Lymphocytes | 8.6 - 47.2 | 36.99 | 2.76 | 17.44 | 1.65 | 11.91 | 1.48 | 25.46 | 2.52 |
| CD3 | 60.3 - 90.6 | 73.30 | 3.01 | 70.94 | 1.54 | 68.33 | 1.98 | 74.61 | 2.50 |
| CD20 | 2.4 - 21.4 | 10.39 | 1.06 | 15.31 | 1.53 | 17.08 | 2.28 | 12.03 | 1.93 |
| CD3-CD16+ or CD56+ | 2.3 - 24.7 | 14.13 | 2.62 | 10.22 | 1.09 | 10.03 | 1.38 | 11.00 | 1.95 |
| %TBNK | | 97.82 | 0.38 | 96.47 | 0.44 | 95.43 | 0.69 | 97.64 | 0.34 |
| CD3+CD16+ or CD56+ | 0.3 - 7.9 | 5.73 | 0.88 | 5.75 | 0.59 | 5.20 | 0.89 | 6.84 | 0.75 |
| T Cell Monitoring and Activation | | | | | | | | | |
| CD3+/CD4+ | 28.3 - 69.4 | 46.52 | 2.65 | 46.08 | 1.61 | 47.63 | 2.35 | 42.89 | 2.10 |
| CD3+/CD8+ | 8.6 - 39.5 | 21.47 | 2.14 | 21.43 | 1.72 | 17.60 | 1.42 | 27.60 | 3.33 |
| CD3+/CD4-/CD8- | 0.4 - 5 | 4.49 | 0.81 | 2.77 | 0.26 | 2.58 | 0.30 | 3.26 | 0.49 |
| CD3/alpha-beta | 56 - 87.6 | 67.37 | 3.17 | 66.60 | 1.52 | 64.33 | 1.89 | 69.67 | 2.65 |
| CC3/gamma-delta | 0.2 - 5.8 | 3.36 | 0.81 | 1.93 | 0.31 | 1.50 | 0.35 | 2.67 | 0.58 |
| DNT/alpha-beta | 0.1 - 1.7 | 1.06 | 0.12 | 0.92 | 0.08 | 1.02 | 0.13 | 0.84 | 0.09 |
| DNT/gamma-delta | 0.2 - 4.3 | 3.24 | 0.79 | 1.57 | 0.26 | 1.21 | 0.29 | 2.20 | 0.47 |
| CD3+/CD8+/CD57+ | 1.2 - 17.2 | 7.31 | 1.73 | 8.72 | 1.30 | 5.60 | 0.98 | 13.39 | 2.58 |
| CD3+/HLA-DR | 1.0 - 9.6 | 1.35 | 0.25 | 3.31 | 0.37 | 3.21 | 0.31 | 3.35 | 0.84 |
| CD3+/CD25 | 11.2 - 53.9 | 12.20 | 1.20 | 13.67 | 0.54 | 14.05 | 0.82 | 12.76 | 0.60 |
| T4/T8 Ratio | 0.71 - 4.3 | 2.50 | 0.25 | 2.80 | 0.29 | 3.25 | 0.44 | 2.04 | 0.30 |
| T Cell Memory | | | | | | | | | |
| CD3/CD4/CD45RO | 9.9 - 37.7 | 26.70 | 1.77 | 32.43 | 1.98 | 29.27 | 2.92 | 35.82 | 2.61 |
| CD3/CD4/CD45RA | 3.4 - 37.9 | 23.68 | 2.19 | 19.79 | 2.37 | 26.12 | 3.38 | 10.87 | 2.30 |
| CD3/CD8/CD45RO | 1.0 - 8.3 | 6.71 | 1.00 | 5.02 | 0.67 | 3.41 | 0.58 | 7.39 | 1.28 |
| CD3/CD8/CD45RA | 2.4 - 23 | 15.09 | 1.77 | 16.22 | 1.54 | 14.88 | 1.79 | 18.73 | 2.90 |
| CD4/CRTH2 | 0.2 - 3.29 | 1.31 | 0.14 | 1.46 | 0.12 | 1.41 | 0.15 | 1.53 | 0.21 |
| CD4/CD45RO/CRTH2 | 0.07 - 2.97 | 1.06 | 0.11 | 1.13 | 0.11 | 1.07 | 0.15 | 1.22 | 0.18 |
| CD8/CRTH2 | <2.43 | 3.67 | 1.19 | 1.72 | 0.35 | 1.45 | 0.40 | 2.07 | 0.67 |
| CD8/CD45RO/CRTH2 | <1.45 | 1.76 | 0.58 | 0.72 | 0.20 | 0.47 | 0.12 | 1.09 | 0.46 |
| B Cell Maturation and Subtypes | | | | | | | | | |
| CD20/CD5 | 0.1 - 4.5 | 0.97 | 0.16 | 0.66 | 0.11 | 0.65 | 0.15 | 0.67 | 0.18 |
| CD20/CD27 | 0.4 - 5.4 | 1.91 | 0.26 | 2.19 | 0.31 | 2.23 | 0.40 | 1.95 | 0.54 |
| CD21 B-cells | 73.2 - 95 | 89.17 | 0.92 | 57.27 | 3.43 | 53.80 | 4.66 | 63.54 | 5.40 |
| CD21 dim B-cells | 3.7 - 21.1 | 8.35 | 0.56 | 34.03 | 2.95 | 35.48 | 3.85 | 29.90 | 4.87 |
| IgG B-cells | 2.5 - 17.4 | 4.80 | 0.40 | 8.21 | 0.71 | 7.67 | 0.78 | 9.17 | 1.42 |
| IgA B-cells | 1.5 - 7.3 | 6.61 | 0.62 | 5.55 | 0.55 | 5.83 | 0.86 | 4.82 | 0.63 |
| IgM B-cells | 69.1 - 97.6 | 85.39 | 0.79 | 84.49 | 0.98 | 84.29 | 1.24 | 84.73 | 1.81 |
| IgD B-cells | 69.0 - 97.6 | 85.17 | 1.08 | 77.97 | 1.39 | 77.42 | 1.54 | 78.48 | 2.81 |
| IgM/CD27 B-cells | 5.6 - 27.2 | 13.60 | 1.43 | 8.76 | 0.84 | 9.02 | 1.17 | 8.79 | 1.36 |
| Dendritic Cell Monitoring | | | | | | | | | |
| DC | 0.04 - 0.5 | 0.17 | 0.02 | 0.13 | 0.03 | 0.09 | 0.02 | 0.19 | 0.06 |
| CD11d | 32.6 - 86 | 60.17 | 3.03 | 60.89 | 4.22 | 50.91 | 5.82 | 72.42 | 5.44 |
| BDCA2 | 8.9 - 59.9 | 31.10 | 3.35 | 9.80 | 1.77 | 8.02 | 2.02 | 13.38 | 3.27 |

| Lymphocyte Monitoring | % CD3- Caspase 1 Nigericin + | % CD3- Caspase 1 Nigericin - | % CD3+ Caspase 1 Nigericin + | % CD3+ Caspase 1 Nigericin - | % CD3+CD4 Caspase 1 Nigericin + | % CD3+CD4 Caspase 1 Nigericin - |
|---|---|---|---|---|---|---|
| Monocytes | 0.0478 | 0.2364 | 0.0034 | 0.0535 | 0.0071 | 0.1902 |
| Lymphocytes | 0.1022 | 0.0938 | 0.4072 | 0.1261 | 0.6053 | 0.5053 |
| CD3 | 0.2239 | 0.0794 | 0.8922 | 0.9462 | 0.0994 | 0.4789 |
| CD20 | 0.0798 | 0.0024 | 0.4962 | 0.0883 | 0.2428 | 0.2798 |
| CD3-CD16+ or CD56+ | 0.2284 | 0.1597 | 0.0585 | 0.0033 | 0.2261 | 0.0014 |
| %TBNK | 0.1822 | 0.8014 | 0.0707 | 0.1595 | 0.2183 | 0.2105 |
| CD3+CD16+ or CD56+ | 0.3994 | 0.2117 | 0.1702 | 0.0738 | 0.601 | 0.2402 |
| T Cell Monitoring and Activation | | | | | | |
| CD3+/CD4+ | 0.6769 | 0.6096 | 0.1232 | 0.3913 | 0.3922 | 0.4624 |
| CD3+/CD8+ | 0.608 | 0.3932 | 0.3267 | 0.6263 | 0.4266 | 0.7696 |
| CD3+/CD4-/CD8- | 0.1454 | 0.1243 | 0.1001 | 0.0153 | 0.7973 | 0.019 |
| CD3/alpha-beta | 0.4105 | 0.1811 | 0.417 | 0.634 | 0.0417 | 0.2596 |
| CC3/gamma-delta | 0.0141 | 0.1792 | 0.0216 | 0.0077 | 0.7028 | 0.0169 |
| DNT/alpha-beta | 0.1929 | 0.5278 | 0.593 | 0.7579 | 0.9569 | 0.6647 |
| DNT/gamma-delta | 0.0486 | 0.1002 | 0.0434 | 0.0039 | 0.7021 | 0.0043 |
| CD3+/CD8+/CD57+ | 0.1636 | 0.5568 | 0.0764 | 0.1449 | 0.6246 | 0.6792 |
| CD3+/HLA-DR | 0.0776 | 0.9094 | 0.2337 | 0.8529 | 0.4494 | 0.4755 |
| CD3+/CD25 | 0.8964 | 0.9711 | 0.5885 | 0.2219 | 0.2188 | 0.5518 |
| T4/T8 Ratio | 0.4452 | 0.7438 | 0.1734 | 0.6016 | 0.5964 | 0.7592 |
| T Cell Memory | | | | | | |
| CD3/CD4/CD45RO | 0.3395 | 0.8005 | 0.0084 | 0.2178 | 0.0001 | 0.1758 |
| CD3/CD4/CD45RA | 0.5221 | 0.9367 | 0.0018 | 0.0958 | 0.004 | 0.1213 |
| CD3/CD8/CD45RO | 0.0242 | 0.1725 | 0.0948 | 0.1103 | 0.5399 | 0.5101 |
| CD3/CD8/CD45RA | 0.0575 | 0.388 | 0.9182 | 0.3908 | 0.3846 | 0.4088 |
| CD4/CRTH2 | 0.7054 | 0.1075 | 0.028 | 0.0371 | 0.0049 | 0.0081 |
| CD4/CD45RO/CRTH2 | 0.6206 | 0.2372 | 0.0173 | 0.03 | 0.0017 | 0.007 |
| CD8/CRTH2 | 0.6995 | 0.7309 | 0.7685 | 0.9211 | 0.672 | 0.963 |
| CD8/CD45RO/CRTH2 | 0.5391 | 0.1153 | 0.8389 | 0.2586 | 0.5109 | 0.2389 |
| B Cell Maturation and Subtypes | | | | | | |
| CD20/CD5 | 0.36 | 0.6784 | 0.8324 | 0.2975 | 0.46 | 0.3127 |
| CD20/CD27 | 0.2057 | 0.0003 | 0.9959 | 0.0648 | 0.0829 | 0.2098 |
| CD21 B-cells | 0.4523 | 0.5262 | 0.9629 | 0.8012 | 0.5335 | 0.6533 |
| CD21 dim B-cells | 0.5599 | 0.7177 | 0.8625 | 0.813 | 0.6934 | 0.9084 |
| IgG B-cells | 0.355 | 0.0271 | 0.3069 | 0.0782 | 0.3498 | 0.107 |
| IgA B-cells | 0.7526 | 0.3071 | 0.5451 | 0.1946 | 0.8896 | 0.3013 |
| IgM B-cells | 0.9812 | 0.2815 | 0.5311 | 0.1143 | 0.8302 | 0.1667 |
| IgD B-cells | 0.4499 | 0.4945 | 0.5855 | 0.2219 | 0.1677 | 0.1857 |
| IgM/CD27 B-cells | 0.0964 | 0.0342 | 0.183 | 0.0677 | 0.9072 | 0.1732 |
| Dendritic Cell Monitoring | | | | | | |
| DC | 0.3404 | 0.5682 | 0.2457 | 0.0183 | 0.9353 | 0.1098 |
| CD11d | 0.113 | 0.6528 | 0.0007 | 0.5627 | 0.0196 | 0.8607 |
| BDCA2 | 0.687 | 0.003 | 0.27 | 0.0051 | 0.4543 | 0.0187 |

*Proposed Journal:* Journal Clinical Investigation Insights / Journal of Clinical Investigation

Supplementary information on antibodies:

The antibodies utilized from Thermo Fisher Scientific were CD56 SB436 [TULY56], CD45 eF506 [HI130], CD3 FITC [SK7], CD16 PE [B73.1], CD8 PerCP-eF710 [SK1] CD14 PE-CY7 [61D3], CD4 APC [SK-3], CD20 APC-eF780 [2H7] CD25 EF450 [CD25-4E3], CD57 FITC [TBo1], TCRγ-δ PE [B1.1], CD4 PerCP-eF710 [SK-3], CD3 PE-CY7 [SK7], TCR αβ APC [IP26], HLA-DR AF700 [LN3], CD8 APC-eF780 [SL1], IgD SB436 [IA6-2], IgA FC Secondary Antibody FITC, IgG FC Secondary Antibody PE, IgM PerCP-eF710 [SA-DA4], CD19 PE-CY7 [SJ25C1], CD27 APC [O323], CD5 FITC [UCHT2], CD21 PE [HB5], CD27 PerCP-eF710 [O323], CD45RA FITC [HI100], CD45RO PerCP-eF710 [UCHL1], CD294 APC [BM16], CD4 AF700 [RPA-T4], CD3 FITC [SK7], CD14 FITC [61D3], CD16 FITC [3G8], CD19 FITC [SJ25-C1], CD20 FITC [2H7], CD56 FITC [TULY56], CD34 FITC [4H11], CD11c PE [3.9], HLA-DR PerCP-EF710 [L243], CD303a APC [201A], CD4 SB600 [SK-3], CD45RA FITC [HI100], CD3 PE-CY7 [SK7], CD8 AF700 [SK1], CCR5 APC [NP-6G4], CD25 APC [BC96], CD317 PE [26F8], IL-6 PE-CY7 [MQ2-13A5], MIP1-β APC [FL34Z3L].

The antibodies utilized from BD Bioscience were HLADR BV480 [G46-6], CD38 PerCP-CY5.5 [HIT2], CD28 APC [CD28.2], CD45 APC H7 [2D1], CD278 BV421 [DX29], CXCR5 PerCP-CY5.5 [RF8B2], CD127 BV480 [HIL-7R-M21], CD45RO PerCP-CY5.5 [UCHL1], CD20 APC-H7 [2H7], CD11b BV421 [ICRF44], CD16 FITC [NKP15], MIP1-α PE [11A3], HLA-DR PerPC-Cy5.5 [L243]. TNF-α BV421 [Mab11] was from Biolegend (San Diego, CA).
